# Incidence, symptoms and medical care for influenza virus and respiratory syncytial virus illnesses among older adults in Eastern China: Findings from the China Ageing Respiratory Infections Study (CARES), 2015-2017

**DOI:** 10.1101/2024.07.03.24309873

**Authors:** Nancy H. L. Leung, Hongjun Zhang, Jun Zhang, Fenyang Tang, Lin Luan, Benfeng Zheng, Guoqing Chen, Changcheng Li, Qigang Dai, Cuiling Xu, Yuyun Chen, Daniel K. W. Chu, Ying Song, Ran Zhang, Lindsay Kim, Rachael Wendlandt, Huachen Zhu, Fiona Havers, Hongjie Yu, Pat Shifflett, Carolyn M. Greene, Benjamin J. Cowling, Mark G. Thompson, A. Danielle Iuliano, China Ageing Respiratory Infections Study (CARES) investigators

## Abstract

**Introduction:** Few studies have examined the incidence of virologic-confirmed influenza virus and respiratory syncytial virus (RSV) infections in community-dwelling older adults.

**Methods:** We enrolled adults aged 60–89 years in Jiangsu Province, China and followed them weekly from December 2015–September 2017 to identify acute respiratory illnesses (ARI), collect illness information and respiratory specimens for laboratory testing.

**Results:** 1,527 adults were enrolled, 0·4% reported ever receiving influenza vaccination. 95 PCR-confirmed influenza ARIs and 22 RSV ARIs were identified, among whom 4–5% required hospitalization. One death associated with RSV ARI while none for influenza ARIs was observed. From December 2015-August 2016, the cumulative incidences of influenza and RSV ARIs were 0·8% (95% CI:0·3–1·4) and 0·5% (95% CI:0·1–1·0), respectively. From September 2016-August 2017, the cumulative incidences were 6·1% (95% CI:4·7–7·7) and 1·0% (95% CI:0·5–1·6); the influenza and RSV ARI-associated hospitalization incidences were 0·3% (95% CI:0–0·8) and 0·1% (95% CI:0–0·2). Feverishness was more common in influenza (55%) than RSV ARIs (30%, p=0·03). Influenza (12·5 days, p=0·02) and RSV ARI symptoms (14·1 days, p=0·15) lasted longer compared to PCR-negative/other ARIs (11·0 days). Antibiotic use was more common for influenza (65%, p=0·02) and RSV (70%, p=0·04) ARIs than other ARIs (51%).

**Conclusions:** We observed a higher incidence of influenza relative to RSV infections among community-dwelling older adults compared to prior studies. Our findings suggest older adults may benefit from receiving influenza and RSV vaccines to reduce the occurrence of illnesses.

## INTRODUCTION

Respiratory virus infections like influenza virus and respiratory syncytial virus (RSV) are major causes of acute hospitalizations and deaths. In temperate climates infections occur mainly during winter, where in tropical and subtropical climates infections may occur year-round.^1^ In older adults, while the potential severity of influenza illness is commonly known,^2^ studies suggested RSV severity may be similar or greater than influenza.^3^ The World Health Organization recommends prioritizing older adults to receive influenza vaccination but uptake remains low in many locations.^4^ More recently RSV vaccines for older adults were approved in some countries.^5^

Vaccination polices require knowledge of influenza and RSV disease burden. Community-based cohort studies^6–10^ can actively identify respiratory illnesses not captured by routine health systems. However, most of these studies have been conducted in households with children^6,11–13^ and in high-income countries.^14^ Fewer were among older adults^15,16^ including those aged ≥80 years, a group vulnerable to severe complications from these infections,^14^ and in low-and-middle income countries.^12,16–18^ To address this gap, we established a community cohort of older adults in Jiangsu Province, China to characterize the incidence, symptom profiles, and medical care of PCR-confirmed influenza virus and RSV illnesses from December 2015–September 2017.^19^

## METHODS

### Study design

We monitored respiratory illnesses in a prospective longitudinal cohort of older adults aged 60– 89 years in Jiangsu Province, China (**Figure 1A**) from December 2015–September 2017 (**Figure 1B**), A detailed cohort description was previously published.^19^ In brief, we screened and enrolled older adults in New District and Xiangcheng in Suzhou and Economic Development Zone (EDZ) and Tinghu in Yancheng, representing urban and semi-urban areas with different socio-economic development levels in eastern China. We recruited older adults in three age groups (60–69, 70– 79, and 80–89 years old) with oversampling among those aged 80–89 years. At enrolment, we collected information on demographics, household size, influenza vaccination history, smoking status, medical history, cognitive functioning, and self-rated health and functioning.^20^ For cognitive functioning, two measures were used: the Mini-Cog (3-item recall) and Standardized Mini-Mental State Examination (SMMSE).^21,22^

**Figure 1.**
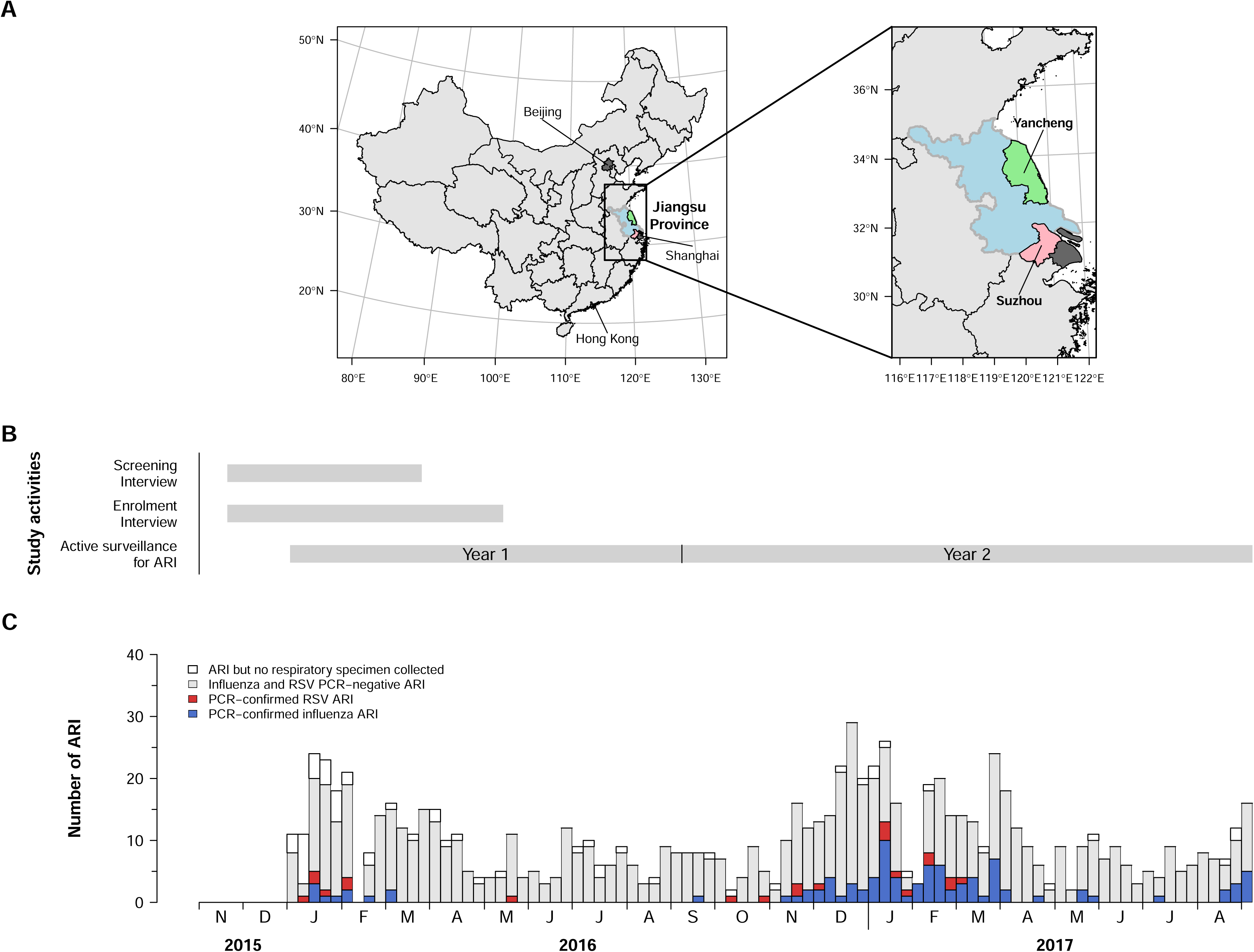
**(A)** CARES study sites – Yancheng and Suzhou, Jiangsu province, mainland China. Blue: Jiangsu province; Green: Yancheng city; Pink: Suzhou city. **(B)** CARES timeline including screening period, enrollment period, and active surveillance for acute respiratory illnesses (ARI). **(C)** Weekly numbers of influenza and RSV PCR-negative ARI, PCR-confirmed influenza ARI, and PCR-confirmed RSV ARI. Blue: PCR-confirmed influenza ARI; Red: PCR-confirmed RSV ARI; Grey: Influenza and RSV PCR-negative ARI; White: ARI but no respiratory specimen collected.

We conducted year-round active surveillance for acute respiratory illness (ARI) (**Appendix Methods**) because influenza viruses are known to circulate beyond the typical winter months in eastern China, and we expect RSV to circulate mostly in winter seasons.^1,23^ During weekly telephone calls, we asked if the participant was experiencing respiratory symptoms. Illness episodes with onset ≥14 days apart were considered separate episodes. Home visits were arranged for new ARI episodes to collect respiratory specimens (combined mid-turbinate and oropharyngeal swabs) within 7 days of symptom onset for laboratory confirmation of infections; study staff also administered an acute illness questionnaire that recorded illness symptoms and whether medical care was sought. Ten days after illness onset, a follow-up questionnaire on illness outcome was completed by telephone. Respiratory specimens were placed in coolers at 2– 8°C and transported to the Suzhou Center for Disease Prevention and Control (CDC) or Yancheng CDC laboratories within 24 hours of collection. Specimens were then stored at -80°C until testing.^19^

### Ethical approval

Written informed consent was obtained from participants. The study was approved by the Institutional Review Board of the University of Hong Kong (UW15 404), the Institutional Review Board of Abt Associates (Abt IRB # 0837) upon which United States CDC relied, and the Ethics Committee of Jiangsu Provincial CDC (JSJK2015-B013-02).

### Laboratory testing

Respiratory specimens were tested for influenza viruses and RSV by real-time reverse transcription-polymerase chain reaction (PCR) using United States CDC primers, probes, reagents, and protocols.^19^

### Primary outcomes and statistical analyses

We measured the cumulative incidence of symptomatic PCR-confirmed influenza virus and RSV infections using the following illness definitions and outcomes: ARI, influenza-like illness (ILI), medically attended ARI, ARI-associated hospitalization, and ARI-associated death (**Appendix Table 1**). An ARI was defined as :22 of the following signs or symptoms with onset in the past 7 days: fever (self-reported feverishness, chills, or temperature ≥37·8°C), cough, sore throat, runny nose or nasal congestion, headache, myalgia (body or muscle aches and pain), or worsened shortness of breath. To compare with prior studies, we examined a subset of ARI as ILI defined as self-reported fever ≥37·8°C plus cough with onset in the past 7 days. A medically attended ARI was defined as an ARI with a self-reported visit to a doctor or other medical professional during the acute illness, including visits to outpatient medical clinic/offices, emergency rooms, or hospitals. An ARI-associated hospitalization was defined as a self-reported hospitalization (i.e., overnight stay in hospital) during the ARI episode. An ARI-associated death was a death that occurred within 14 days of respiratory illness onset. A PCR-positive respiratory specimen indicated a virologic-confirmed influenza virus or RSV infection.

**Table 1.**
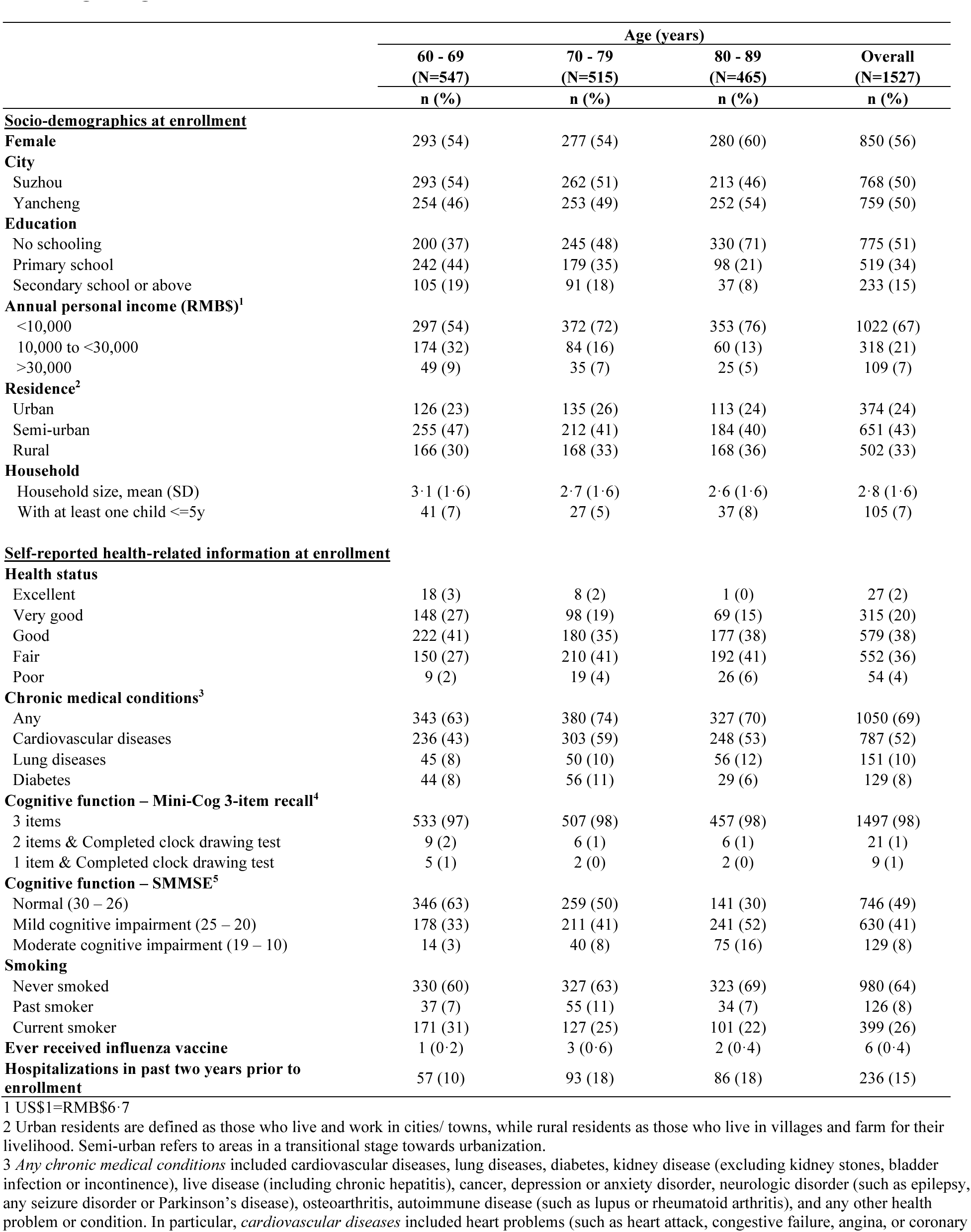
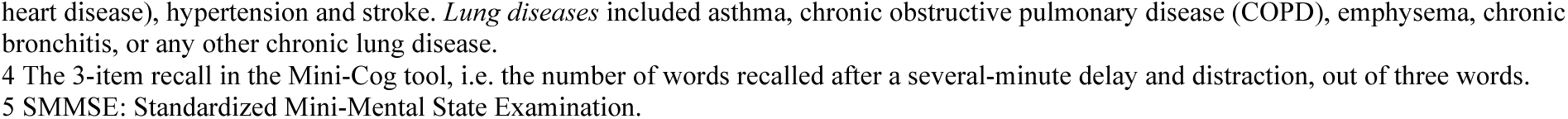
Demographic characteristics at enrolment of an older adult cohort in the cities of Suzhou and Yancheng, Jiangsu Province, China.

We defined the cumulative incidence of influenza or RSV ARI as the proportion of participants with PCR-confirmed influenza virus or RSV infections among participants enrolled at the start of each study year. Study Year 1 was a partial season capturing information from 27 December 2015 through 3 September 2016, and Year 2 was a complete season from 4 September 2016 through 2 September 2017. From previous studies, we anticipated an annual cumulative incidence of influenza ARI of 2–4% in older adults.^12,17^ A sample size of 1,500 participants permitted estimating a cumulative incidence of influenza ARI of 2% with ±0·7% margin of error.^19^ We estimated the cumulative and weekly incidence for each age group, and calculated all-age estimates weighted with the population age and sex structure of Jiangsu Province (**Appendix Figure 1**). We estimated 95% confidence intervals (95% CIs) for the cumulative and weekly incidence using a bootstrap with 2,000 resamples.^24^ If there were no illness outcomes observed during a surveillance week, the point estimate and uncertainty for the true incidence was represented as 0 (0, 3/n) where n was the number of participants followed.^24^

Subjective severity of illness was measured in three ways using previously published methods.^25,26^ First, participants rated the severity of each symptom as absent, mild, moderate, or severe (0–3, respectively), and then a mean symptom severity score (1-3) was calculated.

Second, self-rated overall health during the illness was rated from 0 (worst) to 100 (best health state). Third, the ability to complete regular activities were rated from 0% (unable) to 100% (able to do all activities and responsibilities). Illness duration was defined as the number of days between dates for ARI symptoms onset and resolution with both dates inclusive. We compared illness characteristics of ARI episodes with PCR-confirmed influenza (‘influenza ARI’) or RSV infections (‘RSV ARI’), with ARI episodes that were influenza and RSV PCR-negative (henceforth PCR-negatives). We used Student’s t-test for comparison of severity scores and durations, and Fisher’s exact test for cognitive functioning, symptoms and medical care.

Analyses were done using R version 4.3.1 (R Foundation for Statistical Computing, Vienna, Austria), and geospatial data from the Global Administrative Areas (GADM) database version 2.8.^27^

## RESULTS

We approached 2,280 older adults and screened 1,914 (84%). Of those, 1,573/1,914 (82%) met inclusion criteria, 1,532/1,573 (97%) were enrolled, and 1,527/1,532 (97%) initiated acute respiratory illness surveillance.^19^ As planned, distribution across participant age groups was similar: 60-69 years (n=547, 36%), 70-79 years (515, 34%), and 80–89 years (465, 30%) (**Table 1**). Most participants lived in urban (24%) or semi-urban (43%) areas and had no formal education (51%) or only attended primary school (34%). At enrollment, most participants described overall health status as excellent, very good, or good (59%) compared with fair or poor (40%). Sixty-nine percent of participants self-reported ≥1 chronic health condition and 15% reported hospitalization in the past two years. Assessing cognitive function, almost all participants (98%) recalled three items, with the remaining recalling 1-2 items and having the ability to complete a clock drawing test.

From 27 December 2015—2 September 2017, 1,337/1,527 (88%) participants completed active surveillance follow-up. Among the 190 (12%) lost to follow-up, 84 (44%) died, 7 (4%) withdrew due to deteriorating health and 81 (43%) withdrew for other reasons (**Appendix Table 2**).

**Table 2.** Proportion of participants with acute respiratory illness (ARI), medically attended ARI, ARI- associated hospitalization and death by age group.

|  | Age (years) |  |  |  |
| --- | --- | --- | --- | --- |
|  | 60 - 69 | 70 - 79 | 80 - 89 | Overall |
|  | (N=547) | (N=515) | (N=465) | (N=1527) |
|  | n (%) | n (%) | n (%) | n (%) |
| **Participants followed<sup>1</sup>** |  |  |  |  |
| **Participants at start each study year** |  |  |  |  |
| Year 1 | 547 (100) | 515 (100) | 465 (100) | 1527 (100) |
| Year 2 | 524 (96) | 500 (97) | 446 (96) | 1470 (96) |
| **<u>Acute respiratory illness and medical care</u>** |  |  |  |  |
| **Acute respiratory illness (ARI)** |  |  |  |  |
| Participants with one ARI episode only | 100 (18) | 111 (22) | 87 (19) | 298 (20) |
| Participants with two or more ARI episodes | 86 (16) | 86 (17) | 52 (11) | 224 (15) |
| Participants with one influenza ARI episode only | 34 (6·2) | 36 (7·0) | 21 (4·5) | 91 (6) |
| Participants with two or more influenza ARI episodes | 1 (0·2) | 1 (0·2) | 0 (0) | 2 (0·1) |
| Participants with one RSV ARI episode only | 8 (1·5) | 8 (1·6) | 6 (1·3) | 22 (1·4) |
| **Medically attended ARI<sup>2</sup>** |  |  |  |  |
| Participants with one medically attended ARI episode only | 93 (17) | 97 (19) | 67 (14) | 257 (17) |
| Participants with two or more medically attended ARI episodes | 53 (10) | 42 (8) | 34 (7) | 129 (8) |
| Participants with one medically attended influenza ARI episode only | 33 (6) | 30 (5·8) | 13 (2·8) | 76 (5) |
| Participants with two or more medically attended influenza ARI episodes | 0 (0) | 1 (0·2) | 0 (0) | 1 (0·1) |
| Participants with one medically attended RSV ARI episode only | 5 (0·9) | 4 (0·8) | 3 (0·6) | 12 (0·8) |
| **ARI-associated hospitalization<sup>3</sup>** |  |  |  |  |
| Participants with one hospitalization only | 5 (0·9) | 8 (1·6) | 13 (2·8) | 26 (1·7) |
| Participants with one hospitalization with influenza ARI only | 3 (0·5) | 1 (0·2) | 0 (0) | 4 (0·3) |
| Participants with one hospitalization with RSV ARI only | 0 (0) | 1 (0·2) | 0 (0) | 1 (0·1) |
| **Death within 14 days of ARI symptom onset<sup>4</sup>** |  |  |  |  |
| Deaths | 0 (0) | 1 (0·2) | 0 (0) | 1 (0·1) |
| Deaths with influenza | 0 (0) | 0 (0) | 0 (0) | 0 (0) |
| Deaths with RSV | 0 (0) | 1 (0·2) | 0 (0) | 1 (0·1) |

1 All participants were enrolled at the start of the study with no replenishment. Year 1: 27 December 2015 - 3 September 2016; Year 2: 4 September 2016 - 2 September 2017.
2 A medically attended ARI episode is defined as an ARI with a self-reported medical visit to a doctor or other medical professional for this illness including outpatient medical clinic/offices, emergency rooms or was hospitalized, reported during the acute illness.
3 An ARI-associated hospitalization is defined as an ARI with a self-reported hospitalization, reported during the acute illness.
4 One death during ARI with PCR-confirmed RSV infection but this case was not hospitalized.

Approximately 30% of follow-up contacts to participants were through a proxy who had daily interaction with the participant. From 29,551 and 29,822 successful contact person-weeks reported from New District and Xiangcheng in Suzhou, and 20,036 and 41,075 person-weeks from EDZ and Tinghu in Yancheng respectively (**Appendix Tables 3 – 6**), were identified 1,043 episodes of acute illness, of which 898 (86%) reported by 522 participants met the ARI definition. We collected respiratory specimens for 840 (94%) ARI episodes, 95 (11%) were PCR-confirmed influenza virus infections (“influenza ARI”) and 22 (3%) were RSV infections (“RSV ARI”), including 4 co-infections. Two participants (0·1%) had two separate influenza ARIs (**Table 2**). We identified 56 (59%) influenza and 16 (73%) RSV ARIs from December— February, compared with 12 (13%) and none during June—September (**Figure 1C**). The temporal pattern of influenza detections in our cohort was consistent with community surveillance data over the same period (**Appendix Figure 2**). There were 26 (1·7%) ARI-associated hospitalizations; four were influenza ARIs and one was an RSV ARI (**Table 2**). One death associated with RSV ARI was observed and zero for influenza ARI (**Table 2**).

**Figure 2.**
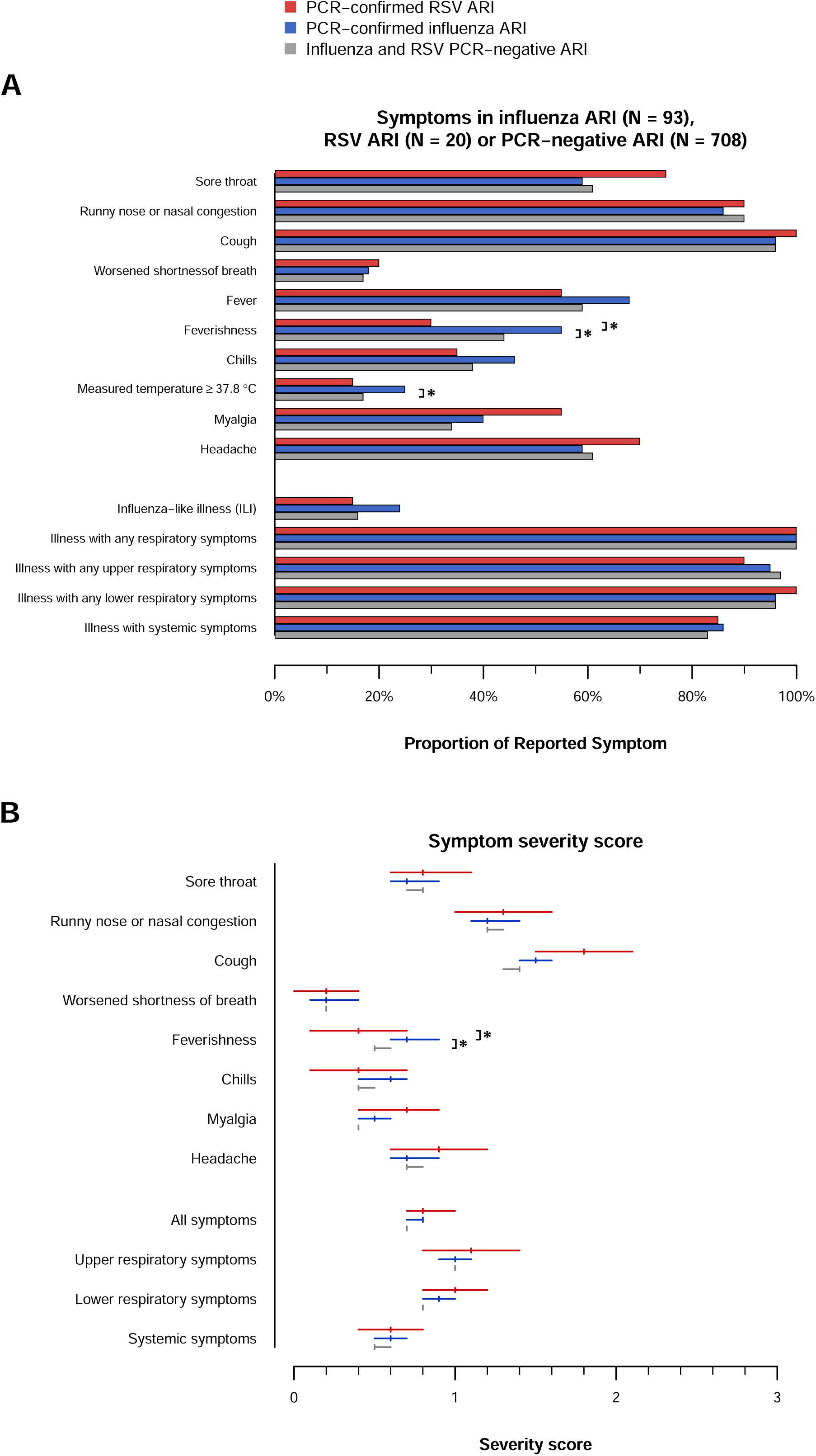
**(A)** Proportion of reported symptoms in participants with PCR-confirmed influenza ARI (N=93), PCR-confirmed RSV ARI (N=20), or influenza and RSV PCR-negative ARI (N=708). **(B)** Mean symptom severity score for reported symptoms and symptom categories (upper respiratory, lower respiratory, or systemic symptoms) in participants with PCR-confirmed influenza ARI, PCR-confirmed RSV ARI, or influenza and RSV PCR-negative ARI. Blue: PCR-confirmed influenza ARI; Red: PCR-confirmed RSV ARI; Grey: Influenza and RSV PCR-negative ARI. * p-value ≤0·05

Of the 95 influenza virus infections, 86 were influenza A, 8 were influenza B, and one was a co-infection with influenza A and B viruses. Influenza A subtyping was completed for 63 specimens; 9 (14%) were A(H1N1) and 54 (86%) were A(H3N2). Twenty-three specimens were not subtyped due to high PCR cycle thresholds. Four influenza B viruses were B/Victoria, three were B/Yamagata, and two were untyped. The most commonly detected virus in Year 1 was A(H1N1) and in Year 2 was A(H3N2) (**Appendix Figure 2**).

We estimated the cumulative incidence (**Table 3**) and incidence rate (**Appendix Table 7**) by age group, and weighted overall estimate, of PCR-confirmed influenza ARI or RSV ARI in each study year. In Year 1, the overall age-weighted incidence of influenza ARI among older adults ≥60 years was 0·8% (95% CI:0·3–1·4), and that of influenza ARI-associated hospitalizations was 0·1% (95% CI:0–0·3). In Year 2, the age-weighted influenza ARI and influenza ARI-associated hospitalization incidences were 6·1% (95% CI:4·7–7·7) and 0·3% (95% CI:0–0·8) respectively. For RSV, in Year 1 the overall age-weighted incidence of RSV ARI was 0·5% (95% CI:0·1–1·0), with no hospitalizations observed. In Year 2, the age-weighted RSV ARI and RSV ARI-associated incidences were 1·0% (95% CI:0·5–1·6) and 0·1% (95% 0–0·2) respectively. Significant differences between age groups were not observed (**Appendix Figure 3**).

**Table 3.** Cumulative incidence of PCR-confirmed influenza ARI, PCR-confirmed RSV ARI, influenza ARI- associated hospitalization and RSV ARI-associated hospitalization.

| Age (years) | Cumulative incidence (%; 95% confidence interval) |  |  |  |
| --- | --- | --- | --- | --- |
|  | 60 - 69 | 70 - 79 | 80 - 89 | Overall (weighted)<sup>6</sup> |
| **Year 1 (27 December 2015 - 3 September 2016)** |  |  |  |  |
| **ARI<sup>1</sup>** | 26·9 (21·9, 32·5) | 23·9 (19·4, 28·7) | 16·1 (12·5, 20·0) | 24·1 (20·7, 27·9) |
| **ILI<sup>2</sup>** | 3·8 (2·0, 5·9) | 3·3 (1·7, 5·2) | 3·4 (1·7, 5·2) | 3·6 (2·4, 5·0) |
| **PCR-confirmed influenza ARI** |  |  |  |  |
| All | 1·1 (0·4, 2·0) | 0·4 (0, 1·0) | 0·4 (0, 1·1) | 0·8 (0·3, 1·4) |
| Medically attended<sup>3</sup> | 1·1 (0·4, 2·0) | 0·2 (0, 0·6) | 0·2 (0, 0·6) | 0·7 (0·2, 1·3) |
| Hospitalized<sup>4</sup> | 0·2 (0, 0·7) | 0 (0, 0·6) | 0 (0, 0·6) | 0·1 (0, 0·3) |
| **PCR-confirmed RSV ARI** |  |  |  |  |
| All | 0·5 (0, 1·3) | 0·6 (0, 1·4) | 0·2 (0, 0·6) | 0·5 (0·1, 1·0) |
| Medically attended | 0·4 (0, 0·9) | 0·2 (0, 0·6) | 0 (0, 0·6) | 0·3 (0, 0·7) |
| Hospitalized<sup>5</sup> | 0 (0, 0·5) | 0 (0, 0·6) | 0 (0, 0·6) | 0 (0, 0·2) |
| **Year 2 (4 September 2016 - 2 September 2017)** |  |  |  |  |
| **ARI** | 38·7 (32·4, 45·4) | 42·0 (35·8, 48·4) | 30·7 (25·3, 37·0) | 39·2 (34·6, 44·5) |
| **ILI** | 5·7 (3·8, 7·8) | 6·8 (4·4, 9·4) | 4·9 (2·9, 7·4) | 5·9 (4·6, 7·5) |
| **PCR-confirmed influenza ARI** |  |  |  |  |
| All | 5·7 (3·8, 7·8) | 7·0 (5·0, 9·4) | 4·3 (2·5, 6·3) | 6·1 (4·7, 7·7) |
| Medically attended | 5·2 (3·4, 7·1) | 6·2 (4·2, 8·6) | 2·7 (1·3, 4·3) | 5·3 (4·1, 6·9) |
| Hospitalized | 0·4 (0, 1·0) | 0·2 (0, 0·6) | 0 (0, 0·7) | 0·3 (0, 0·8) |
| **PCR-confirmed RSV ARI** |  |  |  |  |
| All | 1·0 (0·2, 1·9) | 1·0 (0·2, 2·0) | 1·1 (0·2, 2·2) | 1·0 (0·5, 1·6) |
| Medically attended | 0·6 (0, 1·3) | 0·6 (0, 1·4) | 0·7 (0, 1·6) | 0·6 (0·2, 1·1) |
| Hospitalized | 0 (0, 0·6) | 0·2 (0, 0·6) | 0 (0, 0·7) | 0·1 (0, 0·2) |

1 Acute respiratory illness (ARI) is defined as an acute onset of $\geq 2$ of the following symptoms in the past 7 days: fever (self-reported feverishness, chills, or temperature $\geq 37\cdot8^{\circ}\text{C}$ ), cough, sore throat, runny nose or nasal congestion, headache, myalgia (body or muscle aches and pain), or worsened shortness of breath.
2 Influenza-like illness (ILI) is defined as an ARI with measured fever of $\geq 37\cdot8^{\circ}\text{C}$ (self-reported and/or measured by study staff) plus cough.
3 Medically attended ARI is defined as an ARI with a self-reported medical visit to a doctor or other medical professional during the acute illness, including visits to outpatient medical clinic/offices, emergency rooms, or hospitals.
4 ARI-associated hospitalization is defined as a self-reported hospitalization (i.e. staying in the hospital for at least one night as a patient) during an ARI.
5 There were no hospitalizations with PCR-confirmed RSV infections.
6 The cumulative incidence for each age group was unweighted, while the overall cumulative incidence was weighted with the population age and sex structure of Jiangsu province, China.

Acute illness surveys were completed for 822 (92%) ARI episodes from 487 participants. Comparing ARIs of different virus aetiologies (**Table 4**, **Figure 2, Appendix Figure 4**), subjective feverishness was more common in influenza ARI (55%) compared to RSV ARI (30%, p=0·03) or PCR-negatives (44%, p=0·04). Influenza ARIs met the ILI case definition (24%, p=0·04) significantly more often than PCR-negative ARIs (16%), but not statistically different for RSV ARIs (15%). The sensitivity of the ILI case definition for influenza ARI detection was 24% (95% CI: 15%, 34%) and 15% (95% CI: 3%, 38%) for RSV ARIs; the specificity was 83% (95% CI: 81%, 87%) and 83% (95% CI: 80%, 86%), respectively. Older adults with influenza ARI reported higher symptom severity scores for subjective fever (**Appendix Table 8**) and worsened shortness of breath (**Appendix Table 9**). There were no differences in symptom severity scores by upper respiratory, lower respiratory, or systemic symptom categories. The mean ratings for overall health or ability to do normal activities during the illness were also similar across aetiologies (**Table 4**). Illness duration for an influenza ARI (12·5 days, 95% CI: 11·4, 13·8) was similar to RSV ARI (14·1 days ,95% CI: 10·9, 18·2), but statistically longer (p=0·02) than PCR-negative ARIs (11·0 days, 95% CI: 10·7, 11·3). The number of days spent in bed for influenza ARI (1·2 days, 95% CI: 0·8, 1·7) was also similar to RSV ARI (1·4 days, 95% CI: 0·5, 2·4), but statistically more (p=0·04) than PCR-negative ARIs (0·7 days, 95% CI: 0·6, 0·8).

**Table 4.**
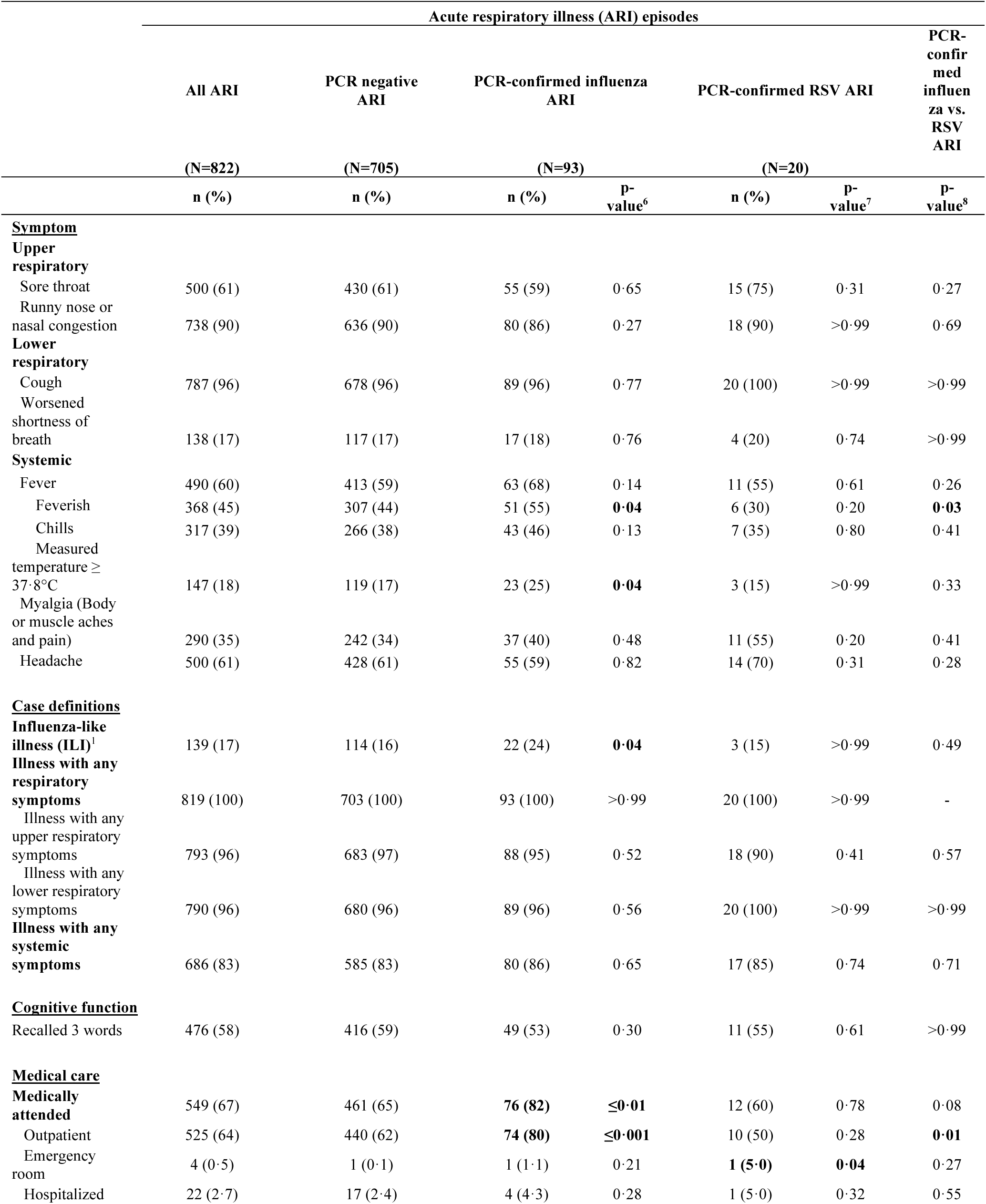
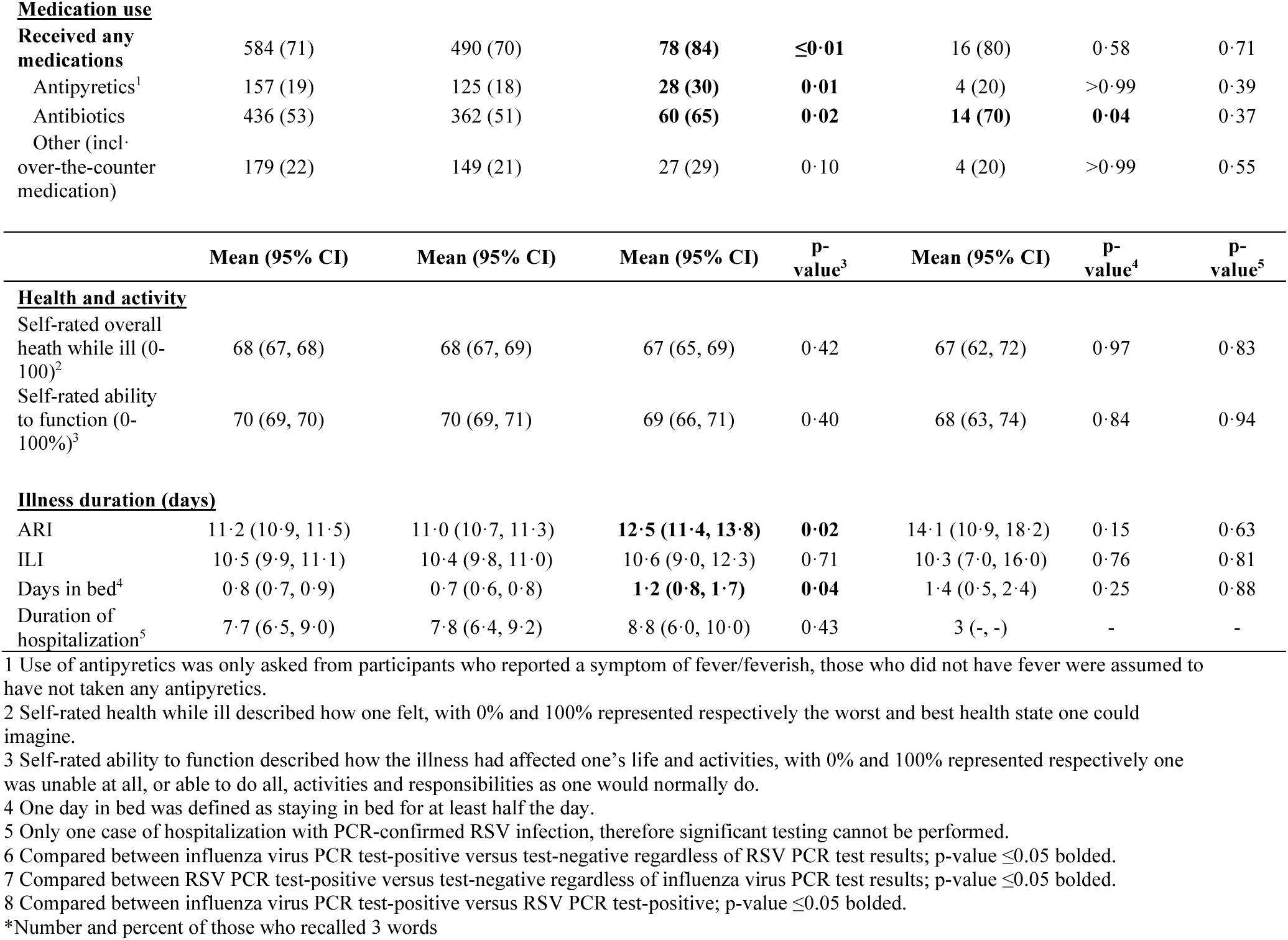
Illness duration, health and functioning, medical care and medication use in PCR-confirmed influenza ARI and RSV ARI.

| Acute respiratory illness (ARI) episodes |  |  |  |  |  |  |  |
| --- | --- | --- | --- | --- | --- | --- | --- |
|  | All ARI | PCR negative ARI | PCR-confirmed influenza ARI |  | PCR-confirmed RSV ARI |  | PCR-confirmed influenza vs. RSV ARI |
|  | (N=822) | (N=705) | (N=93) |  | (N=20) |  |  |
|  | n (%) | n (%) | n (%) | P-value<sup>6</sup> | n (%) | P-value<sup>7</sup> | P-value<sup>8</sup> |
| **Symptom** |  |  |  |  |  |  |  |
| **Upper respiratory** |  |  |  |  |  |  |  |
| Sore throat | 500 (61) | 430 (61) | 55 (59) | 0·65 | 15 (75) | 0·31 | 0·27 |
| Runny nose or nasal congestion | 738 (90) | 636 (90) | 80 (86) | 0·27 | 18 (90) | >0·99 | 0·69 |
| **Lower respiratory** |  |  |  |  |  |  |  |
| Cough | 787 (96) | 678 (96) | 89 (96) | 0·77 | 20 (100) | >0·99 | >0·99 |
| Worsened shortness of breath | 138 (17) | 117 (17) | 17 (18) | 0·76 | 4 (20) | 0·74 | >0·99 |
| **Systemic** |  |  |  |  |  |  |  |
| Fever | 490 (60) | 413 (59) | 63 (68) | 0·14 | 11 (55) | 0·61 | 0·26 |
| Feverish | 368 (45) | 307 (44) | 51 (55) | **0·04** | 6 (30) | 0·20 | **0·03** |
| Chills | 317 (39) | 266 (38) | 43 (46) | 0·13 | 7 (35) | 0·80 | 0·41 |
| Measured temperature ≥ 37·8°C | 147 (18) | 119 (17) | 23 (25) | **0·04** | 3 (15) | >0·99 | 0·33 |
| Myalgia (Body or muscle aches and pain) | 290 (35) | 242 (34) | 37 (40) | 0·48 | 11 (55) | 0·20 | 0·41 |
| Headache | 500 (61) | 428 (61) | 55 (59) | 0·82 | 14 (70) | 0·31 | 0·28 |
| **Case definitions** |  |  |  |  |  |  |  |
| Influenza-like illness (ILI)<sup>1</sup> | 139 (17) | 114 (16) | 22 (24) | **0·04** | 3 (15) | >0·99 | 0·49 |
| Illness with any respiratory symptoms | 819 (100) | 703 (100) | 93 (100) | >0·99 | 20 (100) | >0·99 | - |
| Illness with any upper respiratory symptoms | 793 (96) | 683 (97) | 88 (95) | 0·52 | 18 (90) | 0·41 | 0·57 |
| Illness with any lower respiratory symptoms | 790 (96) | 680 (96) | 89 (96) | 0·56 | 20 (100) | >0·99 | >0·99 |
| Illness with any systemic symptoms | 686 (83) | 585 (83) | 80 (86) | 0·65 | 17 (85) | 0·74 | 0·71 |
| **Cognitive function** |  |  |  |  |  |  |  |
| Recalled 3 words | 476 (58) | 416 (59) | 49 (53) | 0·30 | 11 (55) | 0·61 | >0·99 |
| **Medical care** |  |  |  |  |  |  |  |
| Medically attended | 549 (67) | 461 (65) | **76 (82)** | **≤0·01** | 12 (60) | 0·78 | 0·08 |
| Outpatient | 525 (64) | 440 (62) | **74 (80)** | **≤0·001** | 10 (50) | 0·28 | **0·01** |
| Emergency room | 4 (0·5) | 1 (0·1) | 1 (1·1) | 0·21 | **1 (5·0)** | **0·04** | 0·27 |
| Hospitalized | 22 (2·7) | 17 (2·4) | 4 (4·3) | 0·28 | 1 (5·0) | 0·32 | 0·55 |
| **Medication use** |  |  |  |  |  |  |  |
| **Received any medications** | 584 (71) | 490 (70) | **78 (84)** | **≤0·01** | 16 (80) | 0·58 | 0·71 |
| Antipyretics<sup>1</sup> | 157 (19) | 125 (18) | **28 (30)** | **0·01** | 4 (20) | >0·99 | 0·39 |
| Antibiotics | 436 (53) | 362 (51) | **60 (65)** | **0·02** | **14 (70)** | **0·04** | 0·37 |
| Other (incl. over-the-counter medication) | 179 (22) | 149 (21) | 27 (29) | 0·10 | 4 (20) | >0·99 | 0·55 |

|  | Mean (95% CI) | Mean (95% CI) | Mean (95% CI) | P-value<sup>3</sup> | Mean (95% CI) | P-value<sup>4</sup> | P-value<sup>5</sup> |
| --- | --- | --- | --- | --- | --- | --- | --- |
| **Health and activity** |  |  |  |  |  |  |  |
| Self-rated overall health while ill (0-100)<sup>2</sup> | 68 (67, 68) | 68 (67, 69) | 67 (65, 69) | 0·42 | 67 (62, 72) | 0·97 | 0·83 |
| Self-rated ability to function (0-100%)<sup>3</sup> | 70 (69, 70) | 70 (69, 71) | 69 (66, 71) | 0·40 | 68 (63, 74) | 0·84 | 0·94 |
| **Illness duration (days)** |  |  |  |  |  |  |  |
| ARI | 11·2 (10·9, 11·5) | 11·0 (10·7, 11·3) | **12·5 (11·4, 13·8)** | **0·02** | 14·1 (10·9, 18·2) | 0·15 | 0·63 |
| ILI | 10·5 (9·9, 11·1) | 10·4 (9·8, 11·0) | 10·6 (9·0, 12·3) | 0·71 | 10·3 (7·0, 16·0) | 0·76 | 0·81 |
| Days in bed<sup>4</sup> | 0·8 (0·7, 0·9) | 0·7 (0·6, 0·8) | **1·2 (0·8, 1·7)** | **0·04** | 1·4 (0·5, 2·4) | 0·25 | 0·88 |
| Duration of hospitalization<sup>5</sup> | 7·7 (6·5, 9·0) | 7·8 (6·4, 9·2) | 8·8 (6·0, 10·0) | 0·43 | 3 (-, -) | - | - |

1 Use of antipyretics was only asked from participants who reported a symptom of fever/feverish, those who did not have fever were assumed to have not taken any antipyretics.
2 Self-rated health while ill described how one felt, with 0% and 100% represented respectively the worst and best health state one could imagine.
3 Self-rated ability to function described how the illness had affected one's life and activities, with 0% and 100% represented respectively one was unable at all, or able to do all, activities and responsibilities as one would normally do.
4 One day in bed was defined as staying in bed for at least half the day.
5 Only one case of hospitalization with PCR-confirmed RSV infection, therefore significant testing cannot be performed.
6 Compared between influenza virus PCR test-positive versus test-negative regardless of RSV PCR test results; p-value ≤0.05 bolded.
7 Compared between RSV PCR test-positive versus test-negative regardless of influenza virus PCR test results; p-value ≤0.05 bolded.
8 Compared between influenza virus PCR test-positive versus RSV PCR test-positive; p-value ≤0.05 bolded.
\*Number and percent of those who recalled 3 words

Participants sought medical attention in 67% (549/822) of ARI episodes (**Table 4**). Ambulatory care (outpatient care or visit to an emergency room) was common for both influenza and RSV ARIs, with outpatient care statistically more likely for influenza ARI (80%) compared to RSV ARI (50%, p=0·01) or PCR-negative ARI (62%, p≤0·001). Participants with influenza ARIs (4·3%, p=0·28) and RSV ARIs (5·0%, p=0·32) were hospitalized more frequently than those with PCR-negative ARIs (2·4%) although the difference was not significant. ARI episodes were classified by the highest level of care obtained, the level of care differed significantly by age groups for influenza ARI (p=0·05) and PCR-negative ARI (p=0·03) (**Appendix Table 10**), with a trend of less hospitalizations in older age groups for influenza ARI and a reverse trend for PCR-negative ARI (**Appendix Figure 5**).

Among all ARIs, 71% reported using any medication, with significantly higher use among influenza ARI (84%) than other ARI (**Table 4**). Antibiotic use was reported in 53% of all ARI episodes, with significantly higher use among influenza ARIs (65%) and RSV ARIs (70%) than PCR-negative ARIs (51%).

## DISCUSSION

We estimated the incidence of influenza and RSV ARI, described symptom profiles, and medical care seeking behavior in >1,500 older adults in eastern China between 2015–2017. This was one of the largest prospective observational studies to actively identify laboratory-confirmed influenza and RSV illnesses in older adults. We observed a higher incidence (6·1%) of PCR-confirmed influenza ARI in 2016–17 (study year 2) when circulation of influenza A(H3N2) virus predominated; and a lower incidence (0·8%) in 2015–16 (year 1), based on only a partial study year of active surveillance when circulation of influenza A(H1N1) virus predominated. We estimated PCR-confirmed RSV ARI incidence ranged from 0·5–1·0% in both study years. While our study was not powered to estimate hospitalization incidences, our findings suggested cumulative incidences of 0·1–0·3% and 0–0·1% for hospitalization during acute influenza ARI and RSV ARI, respectively in community-dwelling older adults ≥60 years; and no hospitalizations were observed in those ≥80 years for either influenza or RSV ARI in our cohort. Understanding the effect of influenza virus and RSV infections on older adults provides an evidence base to inform prevention and control measures, including vaccine programs.

Community-based studies of influenza incidence have mostly focused on all ages^12,17,18^ and few focus on older adults specifically.^15^ Tinoco et al. identified ILI, defined as fever or feverishness plus cough or sore throat, by three times weekly home visits in about 500 older adults ≥65 years old from four sites in Peru between 2009–15, with reported influenza vaccination coverage of 26 per 100 person-years.^17^ Prasert et al. identified ARI, defined as new onset of cough or worsening of chronic cough regardless of fever, by weekly calls or home visits in about 1,500 unvaccinated adults ≥65 years in a prospective observational cohort on influenza vaccine effectiveness in Thailand between 2015–17.^15^ Cohen et al. identified infection by nasopharyngeal sampling twice weekly irrespective of symptoms in about 50 older adults ≥65 years (among 1116 participants of all ages from 225 households with almost no influenza vaccination) in South Africa between 2016–18.^18^ Compared to these unvaccinated populations (except Peru), in our study of 1500 older adults ≥60 years, the estimated A(H1N1) ARI incidence (of 1·3/100 person-years) in study year 1 (a partial season) was less than the incidence (based on symptomatic and asymptomatic infections combined) estimated in South Africa (2·3/100 person-years during 2017–18)^18^ and higher than influenza illness in Thailand (<0·3/100 person-years in 2015-16 and 2016-17 seasons).^15^ In study year 2, our estimated A(H3N2) incidence (of 6·5/100 person-years) was higher than Peru (3·6/100 person-years)^17^ and Thailand (<2/100 person-years),^15^ but lower than South Africa (13·6/100 person-seasons during 2017–18).^18^ Thus, our virologic-confirmed influenza incidence estimates fell within the range previously described for community-dwelling older adults, and on the higher end when A(H3N2) viruses predominated. Our study also provided valuable data on influenza ARI-associated hospitalizations in community-dwelling older adults aged ≥80 years, that despite actively identifying ARI we did not observe more influenza ARI- or RSV ARI- associated (acute) hospitalizations in them than those younger between 60-79 years old, although our study was underpowered to observe statistical differences between age groups.

With RSV vaccines now available for older adults, data on RSV incidence in community-dwelling older adults can inform population impact and models evaluating the potential benefits of RSV vaccination programs in older adults, although few of these studies exist, particularly before the COVID-19 pandemic.^6,8,16,28,29^ Prior studies identified RSV illness by passive surveillance only^29^ or evaluated incidence based on both virologic and serologic confirmed infection.^8^ Korsten et al. (RESCEU study) assessed annual RSV community burden in about 500 older adults :260 years old from three cities in Europe between 2017–19 by weekly emails or calls to identify ARI, which was defined as having at least one of the following symptoms: cough, nasal congestion or discharge, wheezing or shortness of breath.^28^ Our study, with active surveillance in >1,500 older adults, found that 0·5–1% experienced virologic-confirmed RSV ARI, 0·3–0·6% had medically attended RSV ARI, and 0–0·1% had RSV ARI-associated hospitalization during 2015–17 in China. In comparison, Korsten et al. reported higher cumulative incidences of virologic-confirmed RSV ARI (2·1% and 4·9% during 2017-18 and 2018-19 October–May seasons respectively) and medically attended RSV ARI (0·8% and 1·4% respectively); and even higher estimates if serologic-confirmed infections.^28^ Our observed lower RSV incidences may be due to differences in surveillance methods, laboratory testing, or true epidemiological variations. Future analyses of our study serum samples^19^ could identify additional RSV infections.

Our study suggested older adults with influenza ARI had longer illnesses than those with PCR-negative ARI, consistent with findings in older adults in RESCEU ^28^ and other working age adults.^26^ We also observed influenza ARI were more likely to include fever compared to PCR-negative ARI or RSV ARI. More influenza ARI met the ILI case definition; and while the sensitivity of the ILI definition to detect either influenza (24%) or RSV (15%) ARI was low, it was relatively higher for influenza ARI, consistent with earlier findings.^10^ This finding suggests the ILI case definition is less effective for identifying RSV in older adults. To capture RSV burden in community-dwelling older adults through routine public health surveillance systems, a new syndromic case definition may be needed.

Over half of our participants with influenza or RSV ARI sought medical care. We observed influenza ARI were more likely to require ambulatory care compared to PCR-negative ARI or RSV ARI, as also reported by Korsten et al.^28^ Influenza vaccine coverage was very low (0·4%) in this population in Jiangsu Province, similar to those in other Chinese cities where influenza vaccines require out-of-pocket payment. Future studies are needed to assess the potential benefit of influenza and RSV vaccination on reducing infections and burden on the healthcare system, and methods that increase influenza vaccine uptake among older adults in China.

Antibiotic use in 9% of all ARI and overuse in 31% of influenza ARI was reported in Europe.^28^ Here, antibiotic use was common in all ARI (53%), particularly for influenza (65%) and RSV (70%) ARI, indicating the broad overuse of antibiotics for non-bacterial infections. Although a decline of antibiotic consumption in health care settings particularly in community care was reported in Shandong Province from 2012–17,^30^ our study suggested antibiotic overuse was still prevalent in the community, and efforts to promote rational use of antibiotics in primary care should be reinforced.

Our study has several limitations. First, there may have been under-detection of illnesses and PCR-confirmed infections, especially in the first study year due to influenza virus circulation before active surveillance began. Second, if a proxy reported on behalf of the participant, some illness episodes may have been missed. However, proxies likely reported most illness episodes as they were required to have daily contact with participants. Finally, we evaluated symptoms and severity within four days of reported illness, potentially missing later-developing symptoms.

Despite these limitations, our study has several important public health implications. We found a higher incidence of PCR-confirmed influenza (mainly from H3N2) in older adults than most previous studies. When the incidence of influenza ARI increased in study year 2, the burden on the healthcare system also increased with more medical care-seeking by older adults, reflecting the effect of seasonal influenza epidemics on healthcare utilisation at the population level. While some cities provide free influenza vaccination for older adults, it is not included in the national immunization program in China and most must pay out-of-pocket. Improving prevention measures such as influenza vaccination in this vulnerable population with extremely low vaccine uptake can reduce infections and healthcare burden. We estimated the incidence of PCR-confirmed RSV illness in older adults in both rural and urban settings in China, providing needed data on RSV infections and burden in older Asian populations to inform the need for RSV vaccination programs. We observed widespread use of antibiotics for viral infections.

## CONTRIBUTORS

All authors meet the International Committee of Medical Journal Editors criteria for authorship. This project is a multi-national collaboration between investigators in the US, Hong Kong, and mainland China, and has required substantial contributions from the co-investigators. In terms of the specific contributions, the study protocol was designed by BJC and MGT and drafted by CX. Substantial input was provided by NHLL, HZ, JZ, FT, LL, BZ, CL, QD, YS, RW, PS, CG, FH, LK, and ADI on the study design in the planning phase in a series of meetings in the US, Hong Kong, and mainland China. The study instruments were prepared by NHLL, CX and RW with input from HZ, JZ, LL, BZ, YC, YS, PS, CG, FH, LK, BJC, MGT and ADI. Study staff training were designed and performed by NHLL, HZ, LL, BZ, QD, CX, YC, YS, RZ, RW, CG and HY. Study activities were managed and organized by HZ, JZ, LL, BZ and YC with oversight by NHLL and BJC. The laboratory tests were developed and performed by GC, DKWC, and HZ. Statistical analyses were done by NHLL. The initial draft of the present manuscript was prepared by NHLL and ADI with input from MGT and BJC, and all authors provided critical review and revision of the text and approved the final version.

## Data Availability

All data produced in the present study are available upon reasonable request to the authors

## ACKNOWLEDGMENTS

CARES investigators also include: Vicky J. Fang, Yi Guan, Chi K. Lam, J. S. Malik Peiris, Yingying Qin (The University of Hong Kong); Min Levine, Brett Whitaker (United States CDC); William Campbell (Abt Associates); Yu Xia, Xuerong Ya, Zefeng Dong (Suzhou CDC); Jinjin Shen, Renjie Jiang, Yao Wang, Yuhong Chen (Yancheng CDC); Yuelong Shu, Luzhao Feng, Jiandong Zheng (China CDC); Weijun Wan (Dongqiao Health Center of Huangdai Town, Xiangcheng District, Suzhou); Hongbo Gu (Healthcare Center of Shishan Street Community, New District, Suzhou); Jinhua Shen (Xiangcheng No. 2 People’s Hospital, Suzhou); Wenjing Wang (Xiangcheng District Center for Disease Prevention and Control, Suzhou); Risheng Zha (New District Center for Disease Prevention and Control, Suzhou); Liwen Gao (Xiangcheng District Health Commission, Suzhou).

The authors would like to thank Eduardo Azziz-Baumgartner, Steve Lindstrom and Jerome Tokars (US CDC) for helpful comments; and Qian Xiong, Hau Chi So, Chi Kin Lam (HKU); Jing Xu (Yancheng CDC); Yuhua Chen, Maogan Wu and Lijuan Wu (Yongfeng Health Center of Tinghu District, Yancheng); Jianhong Wang (Bufeng Health Center of the Economic Development Zone, Yancheng); Ran An (Department of Clinical Laboratory, Yancheng CDC); and other healthcare personnel and colleagues from the Department of Clinical Laboratory, Suzhou CDC; Dongqiao Health Center of Huangdai Town, Xiangcheng District, Suzhou; Healthcare Center of Shishan Street Community, New District, Suzhou; Xiangcheng No. 2 People’s Hospital, Suzhou; Xiangcheng District CDC, Suzhou; New District CDC, Suzhou; and Xiangcheng District Health Commission, Suzhou for technical support in the study.

The findings and conclusions in this report are those of the authors and do not necessarily represent the official position of the U.S. Centers for Disease Control and Prevention or the U.S. Public Health Service.

## REFERENCES

1. Moriyama M, Hugentobler WJ, Iwasaki A. Seasonality of Respiratory Viral Infections. Annu Rev Virol 2020; 7(1): 83–101.

2. Iuliano AD, Roguski KM, Chang HH, et al. Estimates of global seasonal influenza-associated respiratory mortality: a modelling study. Lancet 2018; 391(10127): 1285–300.

3. Ackerson B, Tseng HF, Sy LS, et al. Severe Morbidity and Mortality Associated With Respiratory Syncytial Virus Versus Influenza Infection in Hospitalized Older Adults. Clin Infect Dis 2019; 69(2): 197–203.

4. Palache A, Oriol-Mathieu V, Fino M, Xydia-Charmanta M, Influenza Vaccine Supply task f. Seasonal influenza vaccine dose distribution in 195 countries (2004-2013): Little progress in estimated global vaccination coverage. Vaccine 2015; 33(42): 5598–605.

5. Mazur NI, Terstappen J, Baral R, et al. Respiratory syncytial virus prevention within reach: the vaccine and monoclonal antibody landscape. Lancet Infect Dis 2023; 23(1): e2–e21.

6. Monto AS, Koopman JS, Bryan ER. The Tecumseh Study of Illness. XIV. Occurrence of respiratory viruses, 1976-1981. Am J Epidemiol 1986; 124(3): 359–67.

7. Nicholson KG, Kent J, Hammersley V, Cancio E. Acute viral infections of upper respiratory tract in elderly people living in the community: comparative, prospective, population based study of disease burden. BMJ 1997; 315(7115): 1060–4.

8. Falsey AR, Hennessey PA, Formica MA, Cox C, Walsh EE. Respiratory syncytial virus infection in elderly and high-risk adults. N Engl J Med 2005; 352(17): 1749–59.

9. Xu C, Liu L, Ren B, et al. Incidence of influenza virus infections confirmed by serology in children and adult in a suburb community, northern China, 2018-2019 influenza season. Influenza Other Respir Viruses 2020; 15(2): 262–9.

10. Geismar C, Nguyen V, Fragaszy E, et al. Symptom profiles of community cases infected by influenza, RSV, rhinovirus, seasonal coronavirus, and SARS-CoV-2 variants of concern. Sci Rep 2023; 13(1).

11. Hall CE, Cooney MK, Fox JP. The Seattle virus watch. IV. Comparative epidemiologic observations of infections with influenza A and B viruses, 1965-1969, in families with young children. Am J Epidemiol 1973; 98(5): 365–80.

12. Fowler KB, Gupta V, Sullender W, et al. Incidence of symptomatic A(H1N1)pdm09 influenza during the pandemic and post-pandemic periods in a rural Indian community. Int J Infect Dis 2013; 17(12): e1182–5.

13. Cowling BJ, Perera RA, Fang VJ, et al. Incidence of influenza virus infections in children in Hong Kong in a 3-year randomized placebo-controlled vaccine study, 2009-2012. Clin Infect Dis 2014; 59(4): 517–24.

14. Nguyen-Van-Tam JS, O’Leary M, Martin ET, et al. Burden of respiratory syncytial virus infection in older and high-risk adults: a systematic review and meta-analysis of the evidence from developed countries. Eur Respir Rev 2022; 31(166).

15. Prasert K, Patumanond J, Praphasiri P, et al. Effectiveness of trivalent inactivated influenza vaccine among community-dwelling older adults in Thailand: A two-year prospective cohort study. Vaccine 2019; 37(6): 783–91.

16. Praphasiri P, Shrestha M, Patumanond J, et al. Underlying cardiopulmonary conditions as a risk factor for influenza and respiratory syncytial virus infection among community-dwelling adults aged ≥ 65 years in Thailand: Findings from a two-year prospective cohort study. Influenza Other Respir Viruses 2021; 15(5): 634–40.

17. Tinoco YO, Azziz-Baumgartner E, Uyeki TM, et al. Burden of Influenza in 4 Ecologically Distinct Regions of Peru: Household Active Surveillance of a Community Cohort, 2009-2015. Clin Infect Dis 2017; 65(9): 1532–41.

18. Cohen C, Kleynhans J, Moyes J, et al. Asymptomatic transmission and high community burden of seasonal influenza in an urban and a rural community in South Africa, 2017-18 (PHIRST): a population cohort study. Lancet Glob Health 2021; 9(6): e863–e74.

19. Cowling BJ, Xu C, Tang F, et al. Cohort profile: the China Ageing REespiratory infections Study (CARES), a prospective cohort study in older adults in Eastern China. BMJ Open 2017; 7(10): e017503.

20. Idler EL, Benyamini Y. Self-rated health and mortality: a review of twenty-seven community studies. J Health Soc Behav 1997; 38(1): 21–37.

21. Borson S, Scanlan J, Brush M, Vitaliano P, Dokmak A. The mini-cog: a cognitive ’vital signs’ measure for dementia screening in multi-lingual elderly. Int J Geriatr Psychiatry 2000; 15(11): 1021–7.

22. Molloy DW, Alemayehu E, Roberts R. Reliability of a Standardized Mini-Mental State Examination compared with the traditional Mini-Mental State Examination. Am J Psychiatry 1991; 148(1): 102–5.

23. Yu H, Alonso WJ, Feng L, et al. Characterization of regional influenza seasonality patterns in China and implications for vaccination strategies: spatio-temporal modeling of surveillance data. PLoS Med 2013; 10(11): e1001552.

24. Hanley JA, Lippman-Hand A. If nothing goes wrong, is everything all right? Interpreting zero numerators. JAMA 1983; 249(13): 1743–5.

25. Sun S, Chen J, Johannesson M, et al. Population health status in China: EQ-5D results, by age, sex and socio-economic status, from the National Health Services Survey 2008. Qual Life Res 2011; 20(3): 309–20.

26. Henkle E, Irving SA, Naleway AL, et al. Comparison of laboratory-confirmed influenza and noninfluenza acute respiratory illness in healthcare personnel during the 2010-2011 influenza season. Infect Control Hosp Epidemiol 2014; 35(5): 538–46.

27. University of California. Global Administrative Areas. 2019. https://gadm.org/ (accessed October 21 2019).

28. Korsten K, Adriaenssens N, Coenen S, et al. Burden of respiratory syncytial virus infection in community-dwelling older adults in Europe (RESCEU): an international prospective cohort study. Eur Respir J 2021; 57(4).

29. Kurai D, Natori M, Yamada M, Zheng R, Saito Y, Takahashi H. Occurrence and disease burden of respiratory syncytial virus and other respiratory pathogens in adults aged ≥65 years in community: A prospective cohort study in Japan. Influenza Other Respir Viruses 2021; 16(2): 298–307.

30. Song Y, Han Z, Song K, Zhen T. Antibiotic Consumption Trends in China: Evidence From Six-Year Surveillance Sales Records in Shandong Province. Front Pharmacol 2020; 11.

