## Appendix for "Incidence, symptoms and medical care for influenza virus and respiratory syncytial virus illnesses among older adults in Eastern China: Findings from the China Ageing Respiratory Infections Study (CARES), 2015-2017"

### Additional information on Methods

***Monitoring for acute respiratory illness (ARI)***

We conducted active surveillance for acute respiratory illness (ARI) throughout the year (December 2015-September 2017) with weekly telephone calls because influenza viruses are known to circulate outside of the typical winter season in eastern China, though we expected RSV to circulate mostly in winter seasons.[1,2] To monitor for ARI, we contacted participants every week by telephone until the participants or their proxies were successfully reached. Follow-up with a proxy was used if the participant did not have a phone or was unreachable at the time that weekly calls were conducted. To allow assessment of the quality of reporting by beginning in May 2016, we recorded whether the participant or a proxy was reached during the weekly follow-up calls for illness. If the study participant was not present during the weekly follow-up call and a proxy answered questions about participant illness, the proxy was asked if they interacted with the participant daily since the last follow-up call. During the weekly calls, we asked if the participant was experiencing a new onset of an acute illness by asking about respiratory symptoms. If fever was reported, the participant was asked whether it was subjective feverishness, chills, or measured temperature ≥37·8°C and the participant was asked about other symptoms being experienced. Illness episodes with onset dates that were ≥14 days apart were considered separate illness episodes. If an ARI episode was identified, a home visit was arranged to collect respiratory swab specimens (combined mid-turbinate and oropharyngeal swab) within 7 days of symptom onset for laboratory confirmation of influenza virus and RSV infections. During the household visit, study staff completed an acute illness episode questionnaire that recorded illness symptoms (such as cough, sore throat, congestion, self-reported fever or measured temperature) and duration, clinical characteristics, self-reported health and functioning, cognitive functioning, and whether medical care was sought. Participants described subjective severity of each symptom as absent, mild, moderate, or severe (0–3, respectively),[3,4] and mean symptom severity score (1-3) was calculated for those meeting the ARI definition. Participants reported their overall health on the day of the acute illness survey using a visual analogue thermometer (from 0 [worst health state you can imagine] to 100 [best health state you can imagine]).[5] During the same survey, participants rated their ability to complete their regular activities from 0% (unable to do any activities and responsibilities) to 100% (able to do all activities and responsibilities). Cognitive functioning was assessed with short-term recall of three words.[6,7] About 10 days after illness onset, a follow-up questionnaire was completed by telephone to obtain information about illness resolution and outcome. Participants were asked if they had recovered at 10 days after illness onset; if not, they were contacted every three days by telephone until the illness resolved or there was confirmation of patient death. Additionally, participants also reported whether or not they received medical care during their illness (including hospitalization), the use of antibiotics or over-the-counter medications, and the number days spent in bed for at least half of the day.[8]

Data were captured and stored in a real-time online database, REDCap (Research Electronic Data Capture).[9] Weekly data checks were completed to ensure that all participants were contacted weekly. Respiratory specimens were placed in coolers at 2–8°C and transported to the Suzhou Center for Disease Prevention and Control (CDC) or Yancheng CDC laboratories within 24 hours of collection. Aliquots of specimens were stored at -80°C until testing was completed.[7]

***Primary outcomes and statistical analyses***

We defined the cumulative incidence of influenza or RSV ARIs as the proportion of participants with PCR-confirmed influenza virus or RSV infections among participants enrolled at the start of each study year. Study Year 1 was 27 December 2015–3 September 2016 and Year 2 was 4 September 2016–2 September 2017. Based on previous studies, we anticipated a cumulative incidence of influenza ARI of 2–4% in older adults per year.[10,11] To estimate a cumulative incidence of influenza ARI of 2% with ±0·7% margin of error, we aimed to enroll a sample size of 1,500 older adults.[7] We also estimated the weekly incidence rates of influenza or RSV viruses from symptom onset date per 10,000 person-weeks of follow-up, defining each week from Sunday through Saturday. Participants were included in person-week count if they were contacted and responded to follow-up questions in that week.[12] We estimated the cumulative and weekly incidence by age group and calculated all-age estimates, using the Jiangsu Province population to weight by age and sex (**Appendix Figure 1**). We estimated 95% confidence intervals (95% CIs) for the cumulative and weekly incidence using a bootstrap with 2,000 resamples.[13,14] If there were no illness outcomes observed (e.g., ARI, ILI, influenza or RSV) during a surveillance week, the incidence point estimate was zero and uncertainty regarding the true incidence was represented as (0, 3/n) where n was the number of participants followed.[14]

Clinical characteristics, illness duration, cognitive functioning, self-reported health and functioning, and medical care for ARI episodes were captured during illness interviews. Participants described subjective severity of each symptom as absent, mild, moderate, or severe and responses were scored as 0–3, respectively for each severity level.[3,4] We classified symptoms into categories: upper respiratory symptoms (sore throat, runny nose or nasal congestion), lower respiratory symptoms (cough, shortness of breath), and systemic symptoms (myalgias, headache, and self-reported fever including feverishness, chills, or measured fever ≥37·8°C).[15] We constructed symptom severity scores for each category by adding self-reported symptom scores ranging from 0-3 depending on severity level and dividing by the number of symptoms in the symptom category. The constructed symptom category severity scores ranged from 0-3. Illness duration was defined as the number of days experiencing any ARI symptoms. Illness days in bed was calculated as the number days spent in bed for at least half of the day.[8] During illness, cognitive functioning was assessed with short-term recall of three words,[6,7] self-rated (overall) health state with a visual analogue thermometer (from 0 [worst health state you can imagine] to 100 [best health state you can imagine]),[5] and whether participants were able to complete their regular activities from 0% (unable to do any of your activities and responsibilities) to 100% (able to do all your activities and responsibilities). Participants also self-reported whether or not they received medical care during their illness, medication use (e.g., antipyretics, antibiotics, and antiviral medications), and if they were hospitalized (including duration of hospitalization). If participant death was reported, causes of death were obtained through medical records, death certificates, or family members. We compared illness characteristics of ARI episodes with PCR-confirmed influenza ('influenza ARI') or ARI episodes with PCR-confirmed RSV infections ('RSV ARI'), with ARI episodes that were influenza and RSV PCR-negative ('influenza and RSV PCR-negative ARI'): we used Student's t test for comparison of severity scores and illness duration variables, and Fisher's exact test for comparison of cognitive functioning, symptoms and medical care.

Analyses were done using R version 4.2.2 (R Foundation for Statistical Computing, Vienna, Austria), with the geospatial data obtained from the Global Administrative Areas (GADM) database version 2·8.[16]

### Additional information on Results

#### *Active surveillance for acute respiratory illness (ARI)*

During active surveillance for ARI, January 2016–August 2017, we contacted participants every week and either spoke to the individual or a proxy to assess new onset of symptoms since our last contact. One thousand five hundred and two participants translated to a possible 124,091 weeks eligible for active surveillance. In May 2016, we began to record if health status was reported by the participant or a proxy. Of 97,914 successful weekly contacts, 69,911 (71%) reached the participant directly and 26,531 (27%) reached a proxy to access health status. When a proxy was reached, they were asked if they had regular contact with the participant. Nearly all proxy contacts (26,363, 99%) reported meeting the participant daily. Using these weekly contacts, we assessed health status for 120,484/124,091 (97%) person-weeks of follow-up. We identified 1,043 reports of acute illness. Of those, 898 (86%) episodes reported by 522 participants met the ARI definition.

### Appendix Tables

**Appendix Table 1. Definition of illness definitions and outcomes and self-reported health status.**

| **Outcome** | **Definition/ Measurement** | **Reference** |
|---|---|---|
| **Illness** | | |
| Acute respiratory illness (ARI) | Sudden onset of two or more of the following symptoms in the past 7 days: fever (self-reported feverishness, chills, or temperature ⩾ 37·8°C), cough, sore throat, runny nose or nasal congestion, headache, myalgia (body or muscle aches and pain), or worsened shortness of breath. | |
| Influenza-like illness (ILI) | An ARI with measured fever of ⩾37·8℃ (self-reported and/or measured by study staff) plus cough. | |
| Medically-attended ARI | An ARI with a self-reported medical visit to a doctor or other medical professional during the acute illness, including visits to outpatient medical clinic/offices, emergency rooms, or hospitals. | |
| ARI-associated hospitalization | A self-reported hospitalization (i.e. staying in the hospital for at least one night as a patient) during an ARI. | |
| ARI-associated death | A death that occurs within 14 days of ARI symptom onset. | |
| **Health status** | | |
| Number of days in bed | Number of days the participant stayed in bed for at least half the day while he/ she was sick. | Searle SD et al, BMC Geriatr 2008; 8:24. |
| Cognitive function | Two measures were used: (1) A 3-item recall in the Mini-Cog tool, i.e. the number of words recalled after a several-minute delay and distraction. (2) The full Standardised Mini-Mental State Examination (SMMSE). | Borson S et al, Int J Geriatr Psychiatry 2000; 15:1021-7; Molloy DW et al, Am J Psychiatry 1991; 148:102-5. |
| Self-rate health state | A visual analogue scale (VAS) adapted from the EQ-5D with a range from 0 to 100, where the participant self-evaluated how he/she is feeling today, with the best health state he/she can imagine as 100 at the top, and the worst health state he/she can imagine as 0 at the bottom. | Wu C et al, PLoS One 2016; 11:e0164334. |
| Self-rated activity level | Participant was asked to self-evaluate how his/ her illness has affected his/her life and activities with a range from 0 to 100, where 100% means he/ she was able to do all his/ her activities and responsibilities as he/ she would normally, and 0% means he/ she was unable to do any of his/ her activities or responsibilities. | |

**Appendix Table 2. Reasons for participant withdrawal or loss to follow-up.**

| Cause of withdrawal | Number of participants (N = 190) |
|---|---|
| **Death** | **84 (44%)** |
| Cancer | 30 (16%) |
| Stroke | 17 (9%) |
| Cardiovascular disease | 17 (9%) |
| Chronic lung disease | 4 (2%) |
| Neurologic disorder (e.g. Alzheimer's disease) | 3 (2%) |
| Accident | 3 (2%) |
| Diabetes | 2 (1%) |
| Others | 8 (4%) |
| **No longer interested in participating** | **81 (43%)** |
| **Relocated** | **14 (7%)** |
| **Deteriorate health** | **7 (4%)** |
| **Lost contact** | **2 (1%)** |
| **Others** | **2 (1%)** |

**Appendix Table 3. Proportion of participants with acute respiratory illness (ARI), medically attended ARI, ARI-associated hospitalization and death by age group in New District, Suzhou.**

| | Age (years) | | | | | | | |
|---|---|---|---|---|---|---|---|---|
| | Year 1 | | | | Year 2 | | | |
| | Age (years) | | | | Age (years) | | | |
| | 60 – 69 | 70 – 79 | 80 – 89 | Total | 60 – 69 | 70 – 79 | 80 – 89 | Total |
| **Number of person-week with follow-up attempt** | 4423 | 4577 | 3753 | 12753 | 6360 | 6454 | 5128 | 17942 |
| **Number of person-week with successful follow up (n, %)** | 4238 (95.8) | 4356 (95.2) | 3572 (95.2) | 12166 (95.4) | 6176 (97.1) | 6233 (96.6) | 4976 (97) | 17385 (96.9) |

| | No. of participants (%) | | | | No. of participants (%) | | | |
|---|---|---|---|---|---|---|---|---|
| | (N=126) | (N=135) | (N=113) | (N=374) | (N=124) | (N=130) | (N=107) | (N=361) |
| **Participants followed[1]** | | | | | | | | |
| **Number of participants at start of year** | 126 | 135 | 113 | 374 | 124 | 130 | 107 | 361 |
| **Acute respiratory illness and medical care** | | | | | | | | |
| **Acute respiratory illness (ARI)** | | | | | | | | |
| Participants with one ARI episode only | 12 (10) | 20 (15) | 15 (13) | 47 (13) | 21 (17) | 19 (15) | 20 (19) | 60 (17) |
| Participants with two or more ARI episodes | 5 (4) | 1 (1) | 3 (3) | 9 (2) | 2 (2) | 4 (3) | 3 (3) | 9 (2) |
| Participants with one influenza ARI episode only | 0 (0) | 0 (0) | 2 (1.8) | 2 (0.5) | 3 (2.4) | 6 (4.6) | 3 (2.8) | 12 (3.3) |
| Participants with two or more influenza ARI episodes | 0 (0) | 0 (0) | 0 (0) | 0 (0) | 0 (0) | 0 (0) | 0 (0) | 0 (0) |
| Participants with one RSV ARI episode only | 0 (0) | 2 (1.5) | 0 (0) | 2 (0.5) | 1 (0.8) | 0 (0) | 2 (1.9) | 3 (0.8) |
| **Medically attended ARI[2]** | | | | | | | | |
| Participants with one medically attended ARI episode only | 5 (4) | 6 (4) | 12 (11) | 23 (6) | 13 (10) | 13 (10) | 11 (10) | 37 (10) |
| Participants with two or more medically attended ARI episodes | 2 (2) | 0 (0) | 0 (0) | 2 (1) | 0 (0) | 1 (1) | 2 (2) | 3 (1) |
| Participants with one medically attended influenza ARI episode only | 0 (0) | 0 (0) | 1 (0.9) | 1 (0.3) | 2 (1.6) | 4 (3.1) | 2 (1.9) | 8 (2.2) |
| Participants with two or more medically attended influenza ARI episodes | 0 (0) | 0 (0) | 0 (0) | 0 (0) | 0 (0) | 0 (0) | 0 (0) | 0 (0) |
| Participants with one medically attended RSV ARI episode only | 0 (0) | 1 (0.7) | 0 (0) | 1 (0.3) | 1 (0.8) | 0 (0) | 2 (1.9) | 3 (0.8) |
| **ARI-associated hospitalization[3]** | | | | | | | | |
| Participants with one hospitalization only | 0 (0) | 0 (0) | 3 (2.7) | 3 (0.8) | 0 (0) | 0 (0) | 3 (2.8) | 3 (0.8) |
| Participants with one hospitalization with influenza ARI only | 0 (0) | 0 (0) | 0 (0) | 0 (0) | 0 (0) | 0 (0) | 0 (0) | 0 (0) |
| Participants with one hospitalization with RSV ARI only | 0 (0) | 0 (0) | 0 (0) | 0 (0) | 0 (0) | 0 (0) | 0 (0) | 0 (0) |
| **Death within 14 days of ARI symptom onset[4]** | | | | | | | | |
| Deaths | 0 (0) | 0 (0) | 0 (0) | 0 (0) | 0 (0) | 0 (0) | 0 (0) | 0 (0) |
| Deaths with influenza | 0 (0) | 0 (0) | 0 (0) | 0 (0) | 0 (0) | 0 (0) | 0 (0) | 0 (0) |
| Deaths with RSV | 0 (0) | 0 (0) | 0 (0) | 0 (0) | 0 (0) | 0 (0) | 0 (0) | 0 (0) |

1 All participants were enrolled at the start of the study with no replenishment. Year 1: 27 December 2015 – 3 September 2016; Year 2: 4 September 2016 – 2 September 2017.
2 A medically attended ARI episode is defined as an ARI with a self-reported medical visit to a doctor or other medical professional for this illness including outpatient medical clinic/offices, emergency rooms or was hospitalized, reported during the acute illness.
3 An ARI-associated hospitalization is defined as an ARI with a self-reported hospitalization, reported during the acute illness.
4 One death during ARI with PCR-confirmed RSV infection but this case was not hospitalized.

**Appendix Table 4. Proportion of participants with acute respiratory illness (ARI), medically attended ARI, ARI-associated hospitalization, and death by age group in Xiangcheng, Suzhou.**

| | Age (years) | | | | | | | |
|---|---|---|---|---|---|---|---|---|
| | Year 1 | | | | Year 2 | | | |
| | Age (years) | | | | Age (years) | | | |
| | 60 – 69 | 70 – 79 | 80 – 89 | Total | 60 – 69 | 70 – 79 | 80 – 89 | Total |
| **Number of person-week with follow-up attempt** | 5582 | 4465 | 3391 | 13438 | 7364 | 6210 | 4449 | 18023 |
| **Number of person-week with successful follow up (n, %)** | 5264 (94.3) | 4087 (91.5) | 2956 (87.2) | 12307 (91.6) | 7164 (97.3) | 6051 (97.4) | 4300 (96.7) | 17515 (97.2) |
| | No. of participants (%) | | | | No. of participants (%) | | | |
| | (N=126) | (N=135) | (N=113) | (N=374) | (N=124) | (N=130) | (N=107) | (N=361) |
| **Participants followed[1]** | | | | | | | | |
| **Number of participants at start of year** | 167 | 127 | 100 | 394 | 152 | 125 | 93 | 370 |
| **Acute respiratory illness and medical care** | | | | | | | | |
| **Acute respiratory illness (ARI)** | | | | | | | | |
| Participants with one ARI episode only | 33 (20) | 23 (18) | 13 (13) | 69 (18) | 26 (17) | 39 (31) | 16 (17) | 81 (22) |
| Participants with two or more ARI episodes | 14 (8) | 9 (7) | 2 (2) | 25 (6) | 21 (14) | 20 (16) | 12 (13) | 53 (14) |
| Participants with one influenza ARI episode only | 4 (2.4) | 2 (1.6) | 0 (0) | 6 (1.5) | 13 (8.6) | 8 (6.4) | 5 (5.4) | 26 (7) |
| Participants with two or more influenza ARI episodes | 0 (0) | 0 (0) | 0 (0) | 0 (0) | 0 (0) | 1 (0.8) | 0 (0) | 1 (0.3) |
| Participants with one RSV ARI episode only | 1 (0.6) | 1 (0.8) | 0 (0) | 2 (0.5) | 1 (0.7) | 3 (2.4) | 2 (2.2) | 6 (1.6) |
| **Medically attended ARI[2]** | | | | | | | | |
| Participants with one medically attended ARI episode only | 25 (15) | 14 (11) | 9 (9) | 48 (12) | 17 (11) | 23 (18) | 11 (12) | 51 (14) |
| Participants with two or more medically attended ARI episodes | 3 (2) | 1 (1) | 1 (1) | 5 (1) | 13 (9) | 8 (6) | 4 (4) | 25 (7) |
| Participants with one medically attended influenza ARI episode only | 4 (2.4) | 1 (0.8) | 0 (0) | 5 (1.3) | 11 (7.2) | 5 (4) | 1 (1.1) | 17 (4.6) |
| Participants with two or more medically attended influenza ARI episodes | 0 (0) | 0 (0) | 0 (0) | 0 (0) | 0 (0) | 1 (0.8) | 0 (0) | 1 (0.3) |
| Participants with one medically attended RSV ARI episode only | 1 (0.6) | 0 (0) | 0 (0) | 1 (0.3) | 0 (0) | 2 (1.6) | 0 (0) | 2 (0.5) |
| **ARI-associated hospitalization[3]** | | | | | | | | |
| Participants with one hospitalization only | 1 (0.6) | 1 (0.8) | 2 (2) | 4 (1) | 1 (0.7) | 4 (3.2) | 3 (3.2) | 8 (2.2) |
| Participants with one hospitalization with influenza ARI only | 1 (0.6) | 0 (0) | 0 (0) | 1 (0.3) | 1 (0.7) | 0 (0) | 0 (0) | 1 (0.3) |
| Participants with one hospitalization with RSV ARI only | 0 (0) | 0 (0) | 0 (0) | 0 (0) | 0 (0) | 1 (0.8) | 0 (0) | 1 (0.3) |
| **Death within 14 days of ARI symptom onset[4]** | | | | | | | | |
| Deaths | 0 (0) | 0 (0) | 0 (0) | 0 (0) | 0 (0) | 0 (0) | 0 (0) | 0 (0) |
| Deaths with influenza | 0 (0) | 0 (0) | 0 (0) | 0 (0) | 0 (0) | 0 (0) | 0 (0) | 0 (0) |
| Deaths with RSV | 0 (0) | 0 (0) | 0 (0) | 0 (0) | 0 (0) | 0 (0) | 0 (0) | 0 (0) |

1 All participants were enrolled at the start of the study with no replenishment. Year 1: 27 December 2015 – 3 September 2016; Year 2: 4 September 2016 – 2 September 2017.
2 A medically attended ARI episode is defined as an ARI with a self-reported medical visit to a doctor or other medical professional for this illness including outpatient medical clinic/offices, emergency rooms or was hospitalized, reported during the acute illness.
3 An ARI-associated hospitalization is defined as an ARI with a self-reported hospitalization, reported during the acute illness.
4 One death during ARI with PCR-confirmed RSV infection but this case was not hospitalized.

**Appendix Table 5. Proportion of participants with acute respiratory illness (ARI), medically attended ARI, ARI-associated hospitalization, and death by age group in Economic Development Zone (EDZ), Yancheng.**

| | Age (years) | | | | | | | |
|---|---|---|---|---|---|---|---|---|
| | Year 1 | | | | Year 2 | | | |
| | Age (years) | | | | Age (years) | | | |
| | 60 – 69 | 70 – 79 | 80 – 89 | Total | 60 – 69 | 70 – 79 | 80 – 89 | Total |
| **Number of person-week with follow-up attempt** | 2798 | 2772 | 2763 | 8333 | 4032 | 3923 | 4019 | 11974 |
| **Number of person-week with successful follow up (n, %)** | 2710 (96.9) | 2683 (96.8) | 2680 (97) | 8073 (96.9) | 4030 (100) | 3917 (99.8) | 4016 (99.9) | 11963 (99.9) |
| | No. of participants (%) | | | | No. of participants (%) | | | |
| | (N=126) | (N=135) | (N=113) | (N=374) | (N=124) | (N=130) | (N=107) | (N=361) |
| **Participants followed[1]** | | | | | | | | |
| **Number of participants at start of year** | 88 | 85 | 84 | 257 | 82 | 79 | 81 | 242 |
| **Acute respiratory illness and medical care** | | | | | | | | |
| **Acute respiratory illness (ARI)** | | | | | | | | |
| Participants with one ARI episode only | 10 (11) | 11 (13) | 5 (6) | 26 (10) | 12 (15) | 23 (29) | 19 (23) | 54 (22) |
| Participants with two or more ARI episodes | 4 (5) | 4 (5) | 2 (2) | 10 (4) | 8 (10) | 10 (13) | 7 (9) | 25 (10) |
| Participants with one influenza ARI episode only | 0 (0) | 0 (0) | 0 (0) | 0 (0) | 4 (4.9) | 9 (11.4) | 8 (9.9) | 21 (8.7) |
| Participants with two or more influenza ARI episodes | 0 (0) | 0 (0) | 0 (0) | 0 (0) | 0 (0) | 0 (0) | 0 (0) | 0 (0) |
| Participants with one RSV ARI episode only | 1 (1.1) | 0 (0) | 0 (0) | 1 (0.4) | 2 (2.4) | 0 (0) | 1 (1.2) | 3 (1.2) |
| **Medically attended ARI[2]** | | | | | | | | |
| Participants with one medically attended ARI episode only | 7 (8) | 8 (9) | 4 (5) | 19 (7) | 13 (16) | 22 (28) | 17 (21) | 52 (21) |
| Participants with two or more medically attended ARI episodes | 2 (2) | 2 (2) | 2 (2) | 6 (2) | 3 (4) | 5 (6) | 4 (5) | 12 (5) |
| Participants with one medically attended influenza ARI episode only | 0 (0) | 0 (0) | 0 (0) | 0 (0) | 4 (4.9) | 9 (11.4) | 7 (8.6) | 20 (8.3) |
| Participants with two or more medically attended influenza ARI episodes | 0 (0) | 0 (0) | 0 (0) | 0 (0) | 0 (0) | 0 (0) | 0 (0) | 0 (0) |
| Participants with one medically attended RSV ARI episode only | 0 (0) | 0 (0) | 0 (0) | 0 (0) | 1 (1.2) | 0 (0) | 1 (1.2) | 2 (0.8) |
| **ARI-associated hospitalization[3]** | | | | | | | | |
| Participants with one hospitalization only | 0 (0) | 2 (2.4) | 0 (0) | 2 (0.8) | 1 (1.2) | 0 (0) | 1 (1.2) | 2 (0.8) |
| Participants with one hospitalization with influenza ARI only | 0 (0) | 0 (0) | 0 (0) | 0 (0) | 1 (1.2) | 0 (0) | 0 (0) | 1 (0.4) |
| Participants with one hospitalization with RSV ARI only | 0 (0) | 0 (0) | 0 (0) | 0 (0) | 0 (0) | 0 (0) | 0 (0) | 0 (0) |
| **Death within 14 days of ARI symptom onset[4]** | | | | | | | | |
| Deaths | 0 (0) | 0 (0) | 0 (0) | 0 (0) | 0 (0) | 0 (0) | 0 (0) | 0 (0) |
| Deaths with influenza | 0 (0) | 0 (0) | 0 (0) | 0 (0) | 0 (0) | 0 (0) | 0 (0) | 0 (0) |
| Deaths with RSV | 0 (0) | 0 (0) | 0 (0) | 0 (0) | 0 (0) | 0 (0) | 0 (0) | 0 (0) |

1 All participants were enrolled at the start of the study with no replenishment. Year 1: 27 December 2015 – 3 September 2016; Year 2: 4 September 2016 – 2 September 2017.
2 A medically attended ARI episode is defined as an ARI with a self-reported medical visit to a doctor or other medical professional for this illness including outpatient medical clinic/offices, emergency rooms or was hospitalized, reported during the acute illness.
3 An ARI-associated hospitalization is defined as an ARI with a self-reported hospitalization, reported during the acute illness.
4 One death during ARI with PCR-confirmed RSV infection but this case was not hospitalized.

**Appendix Table 6. Proportion of participants with acute respiratory illness (ARI), medically attended ARI, ARI-associated hospitalization, and death by age group in Tinghu, Yancheng.**

| | Age (years) | | | | | | | |
|---|---|---|---|---|---|---|---|---|
| | **Year 1** | | | | **Year 2** | | | |
| | **Age (years)** | | | | **Age (years)** | | | |
| | **60 – 69** | **70 – 79** | **80 – 89** | **Total** | **60 – 69** | **70 – 79** | **80 – 89** | **Total** |
| **Number of person-week with follow-up attempt** | 5596 | 5611 | 5613 | 16820 | 8498 | 8278 | 8032 | 24808 |
| **Number of person-week with successful follow up (n, %)** | 5419 (96.8) | 5423 (96.6) | 5437 (96.9) | 16279 (96.8) | 8494 (100) | 8272 (99.9) | 8030 (100) | 24796 (100) |
| | **No. of participants (%)** | | | | **No. of participants (%)** | | | |
| | **(N=126)** | **(N=135)** | **(N=113)** | **(N=374)** | **(N=124)** | **(N=130)** | **(N=107)** | **(N=361)** |
| **Participants followed[1]** | | | | | | | | |
| **Number of participants at start of year** | 166 | 168 | 168 | 502 | 166 | 166 | 165 | 497 |
| **Acute respiratory illness and medical care** | | | | | | | | |
| **Acute respiratory illness (ARI)** | | | | | | | | |
| Participants with one ARI episode only | 20 (12) | 22 (13) | 23 (14) | 65 (13) | 30 (18) | 29 (17) | 22 (13) | 81 (16) |
| Participants with two or more ARI episodes | 8 (5) | 7 (4) | 2 (1) | 17 (3) | 16 (10) | 11 (7) | 6 (4) | 33 (7) |
| Participants with one influenza ARI episode only | 2 (1.2) | 0 (0) | 0 (0) | 2 (0.4) | 10 (6) | 11 (6.6) | 3 (1.8) | 24 (4.8) |
| Participants with two or more influenza ARI episodes | 0 (0) | 0 (0) | 0 (0) | 0 (0) | 0 (0) | 0 (0) | 0 (0) | 0 (0) |
| Participants with one RSV ARI episode only | 1 (0.6) | 0 (0) | 1 (0.6) | 2 (0.4) | 1 (0.6) | 2 (1.2) | 0 (0) | 3 (0.6) |
| **Medically attended ARI[2]** | | | | | | | | |
| Participants with one medically attended ARI episode only | 19 (11) | 15 (9) | 17 (10) | 51 (10) | 31 (19) | 28 (17) | 23 (14) | 82 (16) |
| Participants with two or more medically attended ARI episodes | 6 (4) | 7 (4) | 1 (1) | 14 (3) | 15 (9) | 11 (7) | 4 (2) | 30 (6) |
| Participants with one medically attended influenza ARI episode only | 2 (1.2) | 0 (0) | 0 (0) | 2 (0.4) | 10 (6) | 11 (6.6) | 2 (1.2) | 23 (4.6) |
| Participants with two or more medically attended influenza ARI episodes | 0 (0) | 0 (0) | 0 (0) | 0 (0) | 0 (0) | 0 (0) | 0 (0) | 0 (0) |
| Participants with one medically attended RSV ARI episode only | 1 (0.6) | 0 (0) | 0 (0) | 1 (0.2) | 1 (0.6) | 1 (0.6) | 0 (0) | 2 (0.4) |
| **ARI-associated hospitalization[3]** | | | | | | | | |
| Participants with one hospitalization only | 0 (0) | 0 (0) | 0 (0) | 0 (0) | 2 (1.2) | 1 (0.6) | 1 (0.6) | 4 (0.8) |
| Participants with one hospitalization with influenza ARI only | 0 (0) | 0 (0) | 0 (0) | 0 (0) | 0 (0) | 1 (0.6) | 0 (0) | 1 (0.2) |
| Participants with one hospitalization with RSV ARI only | 0 (0) | 0 (0) | 0 (0) | 0 (0) | 0 (0) | 0 (0) | 0 (0) | 0 (0) |
| **Death within 14 days of ARI symptom onset[4]** | | | | | | | | |
| Deaths | 0 (0) | 0 (0) | 0 (0) | 0 (0) | 0 (0) | 1 (0.6) | 0 (0) | 1 (0.2) |
| Deaths with influenza | 0 (0) | 0 (0) | 0 (0) | 0 (0) | 0 (0) | 0 (0) | 0 (0) | 0 (0) |
| Deaths with RSV | 0 (0) | 0 (0) | 0 (0) | 0 (0) | 0 (0) | 1 (0.6) | 0 (0) | 1 (0.2) |

1 All participants were enrolled at the start of the study with no replenishment. Year 1: 27 December 2015 – 3 September 2016; Year 2: 4 September 2016 – 2 September 2017.
2 A medically attended ARI episode is defined as an ARI with a self-reported medical visit to a doctor or other medical professional for this illness including outpatient medical clinic/offices, emergency rooms or was hospitalized, reported during the acute illness.
3 An ARI-associated hospitalization is defined as an ARI with a self-reported hospitalization, reported during the acute illness.
4 One death during ARI with PCR-confirmed RSV infection but this case was not hospitalized.

**Appendix Table 7. Incidence rate of acute respiratory illness (ARI), influenza-like illness (ILI), PCR-confirmed influenza ARI, and PCR-confirmed RSV ARI per 10,000 person-years by age group.**

| | Incidence rate per 10,000 person-years (%, 95% confidence interval) | | | |
|---|---|---|---|---|
| **Age (years)** | **60 – 69** | **70 – 79** | **80 – 89** | **Overall (weighed)[6]** |
| **<u>Year 1</u> (27 December 2015 – 3 September 2016)** | | | | |
| **Number of person-week with follow-up attempt** | 18399 | 17425 | 15520 | 51344 |
| **Number of person-week with successful follow up (n, %)** | 17631 (95·8%) | 16549 (95·0%) | 14645 (94·4%) | 48825 (95·1%) |
| **Acute respiratory illness (ARI)[1]** | 4380 (3699, 5061) | 3878 (3248, 4572) | 2672 (2102, 3243) | 3940 (3473, 4394) |
| **Influenza-like illness (ILI)[2]** | 621 (385, 888) | 536 (315, 788) | 570 (321, 856) | 590 (422, 789) |
| **PCR-confirmed influenza ARI** | | | | |
| All | 178 (59, 355) | 63 (0, 158) | 71 (0, 178) | 125 (49, 220) |
| Medically attended[3] | 178 (59, 326) | 32 (0, 95) | 36 (0, 107) | 111 (34, 212) |
| Hospitalized[4] | 30 (0, 89) | 0 (0, 95) | 0 (0, 107) | 15 (0, 44) |
| **PCR-confirmed RSV ARI** | | | | |
| All | 89 (0, 207) | 95 (0, 221) | 36 (0, 107) | 83 (26, 172) |
| Medically attended | 59 (0, 148) | 32 (0, 95) | 0 (0, 107) | 42 (0, 116) |
| Hospitalized[5] | 0 (0, 89) | 0 (0, 95) | 0 (0, 107) | 0 (0, 32) |
| **<u>Year 2</u> (4 September 2016 – 2 September 2017)** | | | | |
| **Number of person-week with follow-up attempt** | 26254 | 24865 | 21628 | 72747 |
| **Number of person-week with successful follow up (n, %)** | 25864 (98·5%) | 24473 (98·4%) | 21322 (98·6%) | 71659 (98·5%) |
| **Acute respiratory illness (ARI)** | 4095 (3551, 4660) | 4499 (3902, 5117) | 3402 (2838, 3940) | 4204 (3806, 4640) |
| **Influenza-like illness (ILI)** | 605 (403, 827) | 725 (490, 981) | 514 (318, 783) | 640 (492, 807) |
| **PCR-confirmed influenza ARI** | | | | |
| All | 605 (403, 827) | 768 (533, 1023) | 465 (269, 685) | 648 (501, 821) |
| Medically attended | 545 (343, 746) | 661 (426, 895) | 294 (147, 465) | 572 (429, 738) |
| Hospitalized | 40 (0, 101) | 21 (0, 64) | 0 (0, 73) | 35 (0, 81) |
| **PCR-confirmed RSV ARI** | | | | |
| All | 101 (20, 202) | 107 (21, 213) | 122 (24, 245) | 101 (47, 165) |
| Medically attended | 61 (0, 141) | 64 (0, 149) | 73 (0, 171) | 58 (21, 114) |
| Hospitalized | 0 (0, 61) | 21 (0, 64) | 0 (0, 73) | 6 (0, 17) |

1 Acute respiratory illness (ARI) is defined as an acute onset of ≥2 of the following symptoms in the past 7 days: fever (self-reported feverishness, chills, or temperature ≥37·8°C), cough, sore throat, runny nose or nasal congestion, headache, myalgia (body or muscle aches and pain), or worsened shortness of breath.
2 Influenza-like illness (ILI) is defined as an ARI with measured fever of ≥37·8°C (self-reported and/or measured by study staff) plus cough.
3 Medically-attended ARI is defined as an ARI with a self-reported medical visit to a doctor or other medical professional during the acute illness, including visits to outpatient medical clinic/offices, emergency rooms, or hospitals.
4 ARI-associated hospitalization is defined as a self-reported hospitalization (i.e. staying in the hospital for at least one night as a patient) during an ARI.
5 There were no hospitalizations with PCR-confirmed RSV infections.
6 Weighed with Jiangsu province population age and sex structure.

**Appendix Table 8. Self-reported symptoms and symptom severity score among all acute respiratory illness (ARI), PCR-negative ARI, PCR-confirmed influenza ARI, and PCR-confirmed RSV ARI.**

| | Acute respiratory illness (ARI) episodes | | | | | | |
|---|---|---|---|---|---|---|---|
| | All ARI (N=822) | ARI with PCR negative for both influenza and RSV (N=705) | PCR-confirmed influenza ARI (N=93) | | PCR-confirmed RSV ARI (N=20) | | PCR-confirmed influenza vs. RSV ARI |
| | n (%) | n (%) | n (%) | p-value[3] | n (%) | p-value[4] | p-value[5] |
| <u>**Symptoms**</u> | | | | | | | |
| **Upper respiratory** | | | | | | | |
| Sore throat | 500 (61) | 430 (61) | 55 (59) | 0·65 | 15 (75) | 0·31 | 0·27 |
| Runny nose or nasal congestion | 738 (90) | 636 (90) | 80 (86) | 0·27 | 18 (90) | >0·99 | 0·69 |
| **Lower respiratory** | | | | | | | |
| Cough | 787 (96) | 678 (96) | 89 (96) | 0·77 | 20 (100) | >0·99 | >0·99 |
| Worsened shortness of breath | 138 (17) | 117 (17) | 17 (18) | 0·76 | 4 (20) | 0·74 | >0·99 |
| **Systemic** | | | | | | | |
| Fever | 490 (60) | 413 (59) | 63 (68) | 0·14 | 11 (55) | 0·61 | 0·26 |
| Feverish | 368 (45) | 307 (44) | 51 (55) | **0·04** | 6 (30) | 0·20 | **0·03** |
| Chills | 317 (39) | 266 (38) | 43 (46) | 0·13 | 7 (35) | 0·80 | 0·41 |
| Measured temperature ≥ 37·8°C | 147 (18) | 119 (17) | 23 (25) | **0·04** | 3 (15) | >0·99 | 0·33 |
| Myalgia (Body or muscle aches and pain) | 290 (35) | 242 (34) | 37 (40) | 0·48 | 11 (55) | 0·20 | 0·41 |
| Headache | 500 (61) | 428 (61) | 55 (59) | 0·82 | 14 (70) | 0·31 | 0·28 |
| <u>**Case definitions**</u> | | | | | | | |
| **Influenza-like illness (ILI)[1]** | 139 (17) | 114 (16) | 22 (24) | **0·04** | 3 (15) | >0·99 | 0·49 |
| **Illness with any respiratory symptoms** | 819 (100) | 703 (100) | 93 (100) | >0·99 | 20 (100) | >0·99 | - |
| Illness with any upper respiratory symptoms | 793 (96) | 683 (97) | 88 (95) | 0·52 | 18 (90) | 0·41 | 0·57 |
| Illness with any lower respiratory symptoms | 790 (96) | 680 (96) | 89 (96) | 0·56 | 20 (100) | >0·99 | >0·99 |
| **Illness with any systemic symptoms** | 686 (83) | 585 (83) | 80 (86) | 0·65 | 17 (85) | 0·74 | 0·71 |
| | Mean (95% CI) | Mean (95% CI) | Mean (95% CI) | p-value[3] | n (%) | p-value[4] | p-value[5] |
| <u>**Symptom severity score**</u> | | | | | | | |
| **Symptoms** | | | | | | | |
| Sore throat | 0·8 (0·7, 0·8) | 0·8 (0·7, 0·8) | 0·7 (0·6, 0·8) | 0·41 | 0·8 (0·6, 1·1) | 0·58 | 0·40 |
| Runny nose or nasal congestion | 1·2 (1·2, 1·3) | 1·2 (1·2, 1·3) | 1·2 (1·1, 1·4) | 0·87 | 1·3 (1·0, 1·6) | 0·64 | 0·61 |
| Cough | 1·4 (1·4, 1·5) | 1·4 (1·3, 1·4) | 1·5 (1·3, 1·6) | 0·31 | 1·8 (1·4, 2·1) | 0·06 | 0·15 |
| Worsened shortness of breath | 0·2 (0·2, 0·2) | 0·2 (0·2, 0·2) | 0·2 (0·1, 0·4) | 0·41 | 0·2 (0, 0·4) | 0·96 | 0·62 |
| Feverishness | 0·5 (0·5, 0·6) | 0·5 (0·5, 0·6) | **0·7 (0·6, 0·9)** | **0·02** | 0·4 (0·1, 0·7) | 0·19 | **0·03** |
| Chills | 0·5 (0·4, 0·5) | 0·4 (0·4, 0·5) | 0·6 (0·4, 0·7) | 0·14 | 0·4 (0·2, 0·7) | 0·64 | 0·29 |
| Myalgia (Body or muscle aches and pain) | 0·4 (0·4, 0·5) | 0·4 (0·4, 0·4) | 0·5 (0·3, 0·6) | 0·32 | 0·7 (0·4, 0·9) | 0·23 | 0·44 |
| Headache | 0·7 (0·7, 0·8) | 0·7 (0·7, 0·8) | 0·7 (0·6, 0·8) | 0·87 | 0·9 (0·6, 1·2) | 0·31 | 0·30 |
| **Symptom categories[2]** | | | | | | | |
| All symptoms | 0·7 (0·7, 0·7) | 0·7 (0·7, 0·7) | 0·8 (0·7, 0·8) | 0·19 | 0·8 (0·7, 1·0) | 0·44 | 0·76 |
| Upper respiratory | 1·0 (1·0, 1·0) | 1·0 (1·0, 1·0) | 1·0 (0·9, 1·1) | 0·52 | 1·1 (0·8, 1·4) | 0·58 | 0·46 |
| Lower respiratory | 0·8 (0·8, 0·8) | 0·8 (0·8, 0·8) | 0·9 (0·8, 1·0) | 0·23 | 1·0 (0·8, 1·2) | 0·12 | 0·37 |
| Systemic | 0·5 (0·5, 0·6) | 0·5 (0·5, 0·6) | 0·6 (0·5, 0·7) | 0·08 | 0·6 (0·4, 0·8) | 0·78 | 0·72 |

1 Influenza-like illness (ILI) is defined as an acute onset of measured fever of ≥37·8℃ plus cough.
2 Use of antipyretics was only asked from participants who reported a symptom of fever/feverish, those who did not have fever were assumed to have not taken any antipyretics.

3 Compared between influenza virus PCR test-positive versus test-negative regardless of RSV PCR test results. p-value $\leq 0{\cdot}05$ is highlighted in bold.

4 Compared between RSV PCR test-positive versus test-negative regardless of influenza virus PCR test results. p-value $\leq 0{\cdot}05$ is highlighted in bold.

5 Compared between influenza virus PCR test-positive versus RSV PCR test-positive. p-value $\leq 0{\cdot}05$ is highlighted in bold.

6 All the information excepted measured body temperature $\geq 37{\cdot}8$°C have <5% unknown status, with unknown status assumed to be absent.

**Appendix Table 9. Self-reported symptom severity score among acute respiratory illness (ARI) episodes presenting the specific symptom.**

| | Acute respiratory illness (ARI) episodes | | | | | | | | | | |
|---|---|---|---|---|---|---|---|---|---|---|---|
| | All ARI (N=822) | | ARI with PCR negative for both influenza and RSV (N=705) | | PCR-confirmed influenza ARI (N=93) | | | PCR-confirmed RSV ARI (N=20) | | | PCR-confirmed influenza vs. RSV ARI |
| | n[1] (%) | Mean (95% CI) | n[1] (%) | Mean (95% CI) | n[1] (%) | Mean (95% CI) | p-value[2] | n[1] (%) | Mean (95% CI) | p-value[3] | p-value[4] |
| **Symptom severity score** | | | | | | | | | | | |
| Sore throat | 500 (61) | 1·3 (1·2, 1·3) | 430 (61) | 1·3 (1·2, 1·3) | 55 (59) | 1·2 (1·1, 1·3) | 0·46 | 15 (75) | 1·1 (1·0, 1·4) | 0·58 | 0·80 |
| Runny nose or nasal congestion | 737 (90) | 1·4 (1·3, 1·4) | 635 (90) | 1·4 (1·3, 1·4) | 80 (86) | 1·4 (1·3, 1·6) | 0·56 | 18 (90) | 1·4 (1·2, 1·7) | 0·81 | 0·99 |
| Cough | 786 (96) | 1·5 (1·4, 1·5) | 677 (96) | 1·4 (1·4, 1·5) | 89 (96) | 1·6 (1·4, 1·7) | 0·21 | 20 (100) | 1·8 (1·5, 2·1) | 0·10 | 0·27 |
| Worsened shortness of breath | 138 (17) | 1·2 (1·1, 1·3) | 117 (17) | 1·2 (1·1, 1·2) | 17 (18) | 1·4 (1·1, 1·6) | 0·20 | 4 (20) | 1·0 (1·0, 1·0) | **≤0·001** | **0·03** |
| Feverishness | 367 (45) | 1·2 (1·2, 1·3) | 306 (43) | 1·2 (1·1, 1·2) | 51 (55) | 1·3 (1·2, 1·4) | 0·24 | 6 (30) | 1·3 (1·0, 1·7) | 0·84 | 0·90 |
| Chills | 317 (39) | 1·2 (1·1, 1·2) | 266 (38) | 1·2 (1·1, 1·2) | 43 (46) | 1·2 (1·1, 1·3) | 0·70 | 7 (35) | 1·1 (1·0, 1·4) | 0·97 | 0·93 |
| Myalgia (Body or muscle aches and pain) | 289 (35) | 1·2 (1·1, 1·2) | 241 (34) | 1·2 (1·1, 1·2) | 37 (40) | 1·2 (1·1, 1·4) | 0·39 | 11 (55) | 1·2 (1·0, 1·5) | 0·59 | 0·94 |
| Headache | 499 (61) | 1·2 (1·2, 1·2) | 427 (61) | 1·2 (1·2, 1·2) | 55 (59) | 1·2 (1·1, 1·3) | 0·95 | 14 (70) | 1·3 (1·0, 1·6) | 0·80 | 0·82 |

1 n represented number of participants who had the symptom and reported symptom severity score for that specific symptom.
2 Compared between influenza virus PCR test-positive versus test-negative regardless of RSV PCR test results. p-value ≤0·05 is highlighted in bold.
3 Compared between RSV PCR test-positive versus test-negative regardless of influenza virus PCR test results. p-value ≤0·05 is highlighted in bold.
4 Compared between influenza virus PCR test-positive versus RSV PCR test-positive. p-value ≤0·05 is highlighted in bold.

**Appendix Table 10. Self-reported medical care sought for PCR-confirmed influenza or RSV ARIs.**

| | Age (years) | | | | | | | |
|---|---|---|---|---|---|---|---|---|
| | 60 - 69 | | 70 - 79 | | 80 - 89 | | Overall | |
| | **n** | **% (95% CI)** | **n** | **% (95% CI)** | **n** | **% (95% CI)** | **n** | **% (95% CI)** |
| **All ARI (N=822)**, p-value = 0·36 | | | | | | | | |
| Hospitalized | 5 | 1·6% (0·3, 3·1) | 8 | 2·6% (1·0, 4·6) | 9 | 4·5% (2·0, 7·6) | 22 | 2·7% (1·6, 3·8) |
| Ambulatory care only | 210 | 65·4% (60·4, 70·4) | 190 | 62·7% (57·1, 68·0) | 127 | 64·1% (57·1, 70·7) | 528 | 64·1% (60·8, 67·3) |
| Did not seek medical attention | 105 | 32·7% (27·7, 37·7) | 103 | 34·0% (28·7, 39·6) | 62 | 31·3% (24·7, 37·9) | 272 | 32·8% (29·7, 36·0) |
| **Influenza and RSV PCR-negative ARI (N=705), p-value = 0·03** | | | | | | | | |
| Hospitalized | 2 | 0·7% (0, 1·8) | 6 | 2·3% (0·8, 4·3) | 9 | 5·3% (2·3, 9·4) | 17 | 2·4% (1·3, 3·7) |
| Ambulatory care only | 176 | 63·5% (57·4, 69·0) | 156 | 60·7% (54·9, 66·5) | 112 | 65·5% (58·5, 71·9) | 445 | 63·0% (59·4, 66·7) |
| Did not seek medical attention | 99 | 35·7% (30·0, 41·5) | 95 | 37·0% (31·1, 43·2) | 50 | 29·2% (22·8, 35·7) | 246 | 34·6% (30·9, 38·0) |
| **PCR-confirmed influenza ARI (N=93), p-value = 0·05** | | | | | | | | |
| Hospitalized | 3 | 8·6% (0, 17·2) | 1 | 2·7% (0, 8·1) | 0 | 0% (0, 3·2) | 4 | 4·3% (1·1, 8·6) |
| Ambulatory care only | 29 | 82·9% (71·4, 94·3) | 30 | 81·1% (67·6, 91·9) | 13 | 61·9% (42·9, 81·0) | 72 | 77·4% (68·8, 86·0) |
| Did not seek medical attention | 3 | 8·6% (0, 20·0) | 6 | 16·2% (5·4, 29·7) | 8 | 38·1% (19·0, 57·1) | 17 | 18·3% (10·8, 26·9) |
| **PCR-confirmed RSV ARI (N=20)**, p-value = 0·74 | | | | | | | | |
| Hospitalized | 0 | 0% (0, 15·0) | 1 | 14·3% (0, 42·9) | 0 | 0% (0, 15·0) | 1 | 5·0% (0, 15·0) |
| Ambulatory care only | 5 | 71·4% (42·9, 100) | 3 | 42·9% (14·3, 85·7) | 3 | 50·0% (16·7, 83·3) | 11 | 55·0% (35·0, 75·0) |
| Did not seek medical attention | 2 | 28·6% (0, 57·1) | 2 | 28·6% (0, 71·4) | 3 | 50·0% (16·7, 83·3) | 7 | 35·0% (15·0, 55·0) |

Note: There were statistically significant differences between age groups and medical care sought, among those with influenza and RSV-negative ARIs (p-value = 0·03), and among those with PCR-confirmed influenza ARIs (p-value = 0·05). Unlike in Table 4, here the categorization to "hospitalized", "ambulatory care only" and "did not seek medical attention" was mutually exclusive.

### Appendix Figures

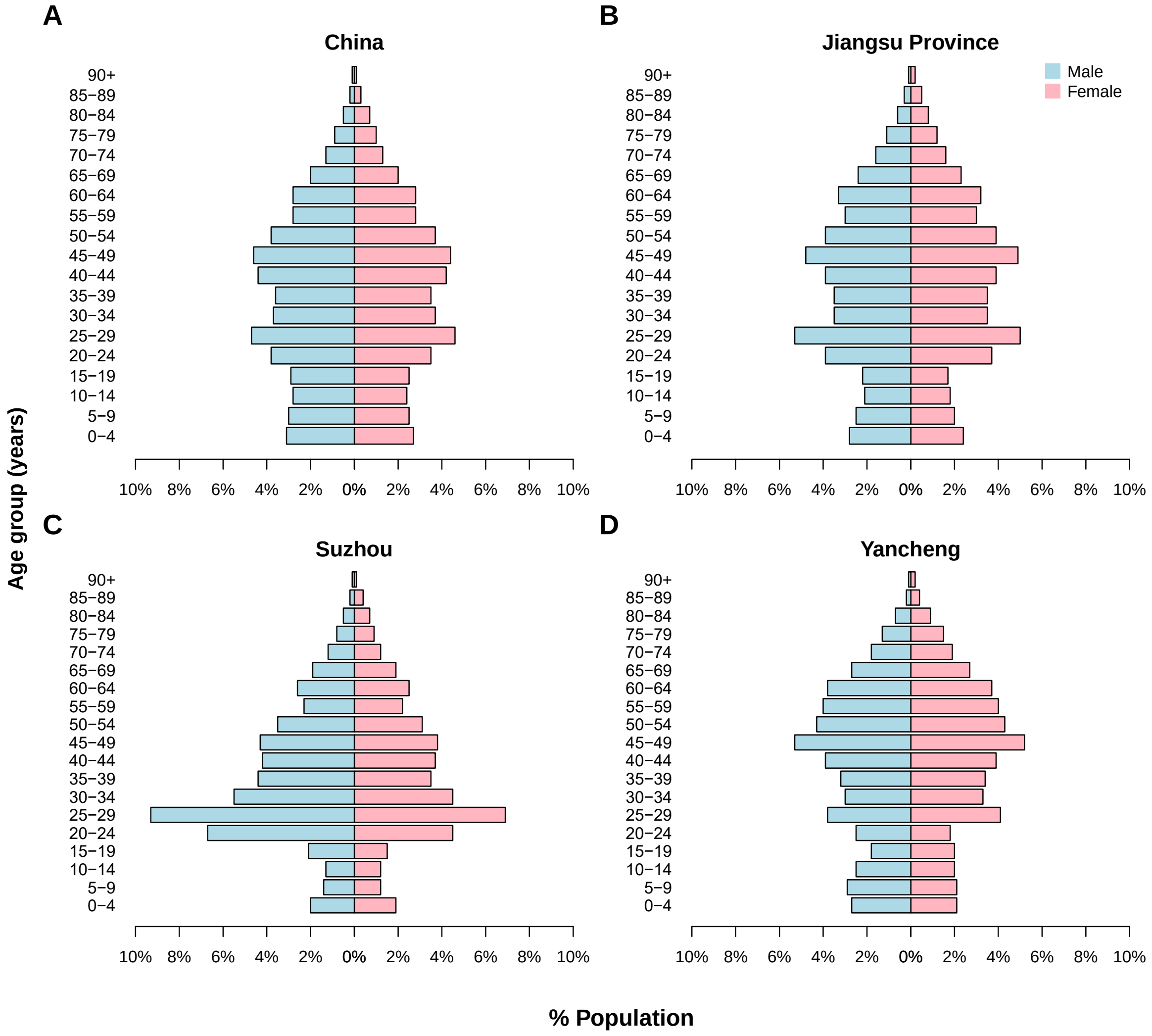

**Appendix Figure 1. 2015 population percentage by five-year age groups and gender for (A) China, (B) Jiangsu Province, (C) Suzhou and (D) Yancheng.**

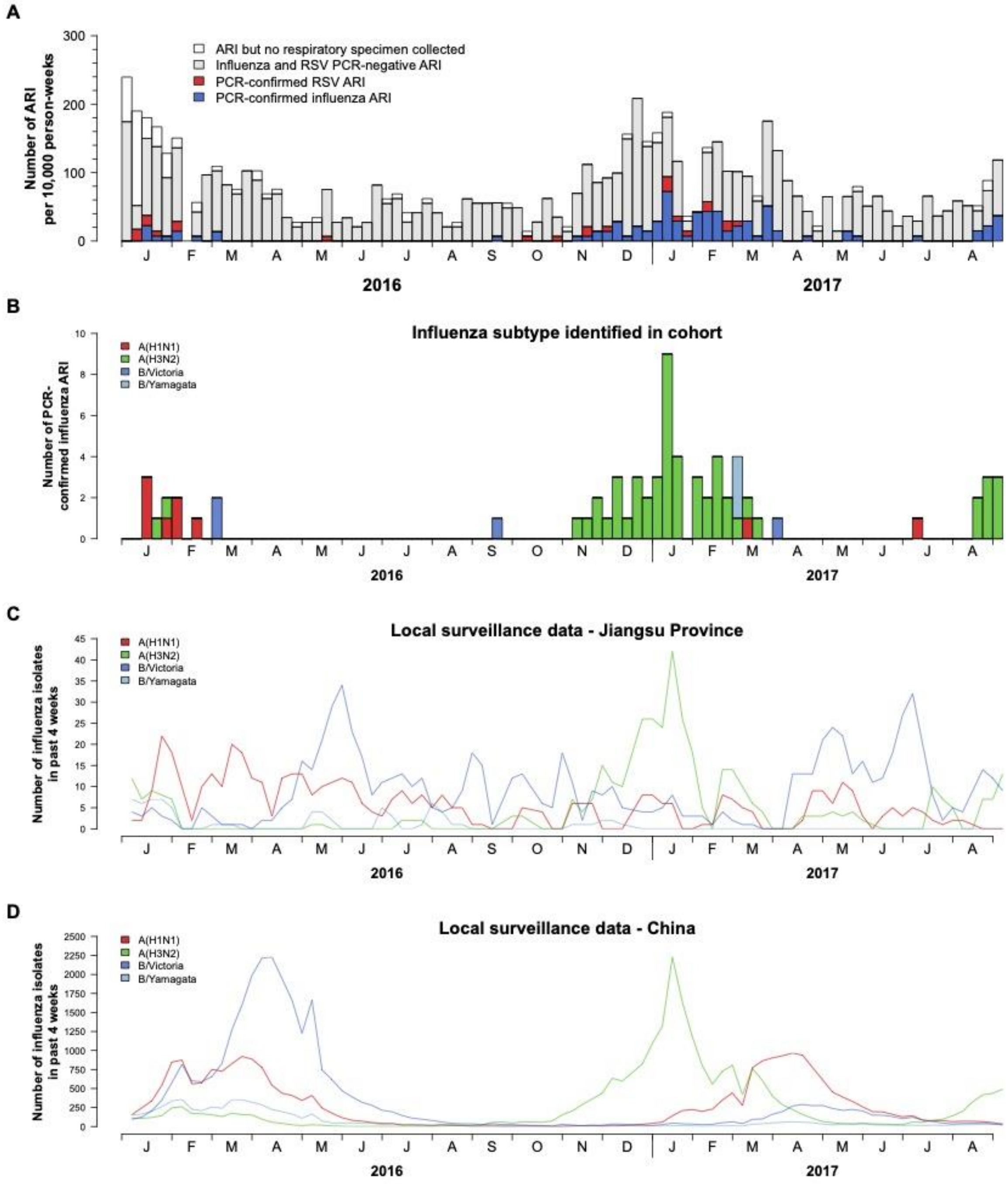

**Appendix Figure 2. (A) Incidence of PCR-confirmed influenza acute respiratory illness (ARI) and RSV ARI per 10,000 person-weeks. (B) Weekly numbers of PCR-confirmed influenza ARI by influenza A virus subtypes or influenza B virus lineages. Number of influenza isolates in the past four weeks in (C) Jiangsu Province and (D) China as reported by Chinese Center for Disease Control and Prevention in Influenza Weekly Report.**

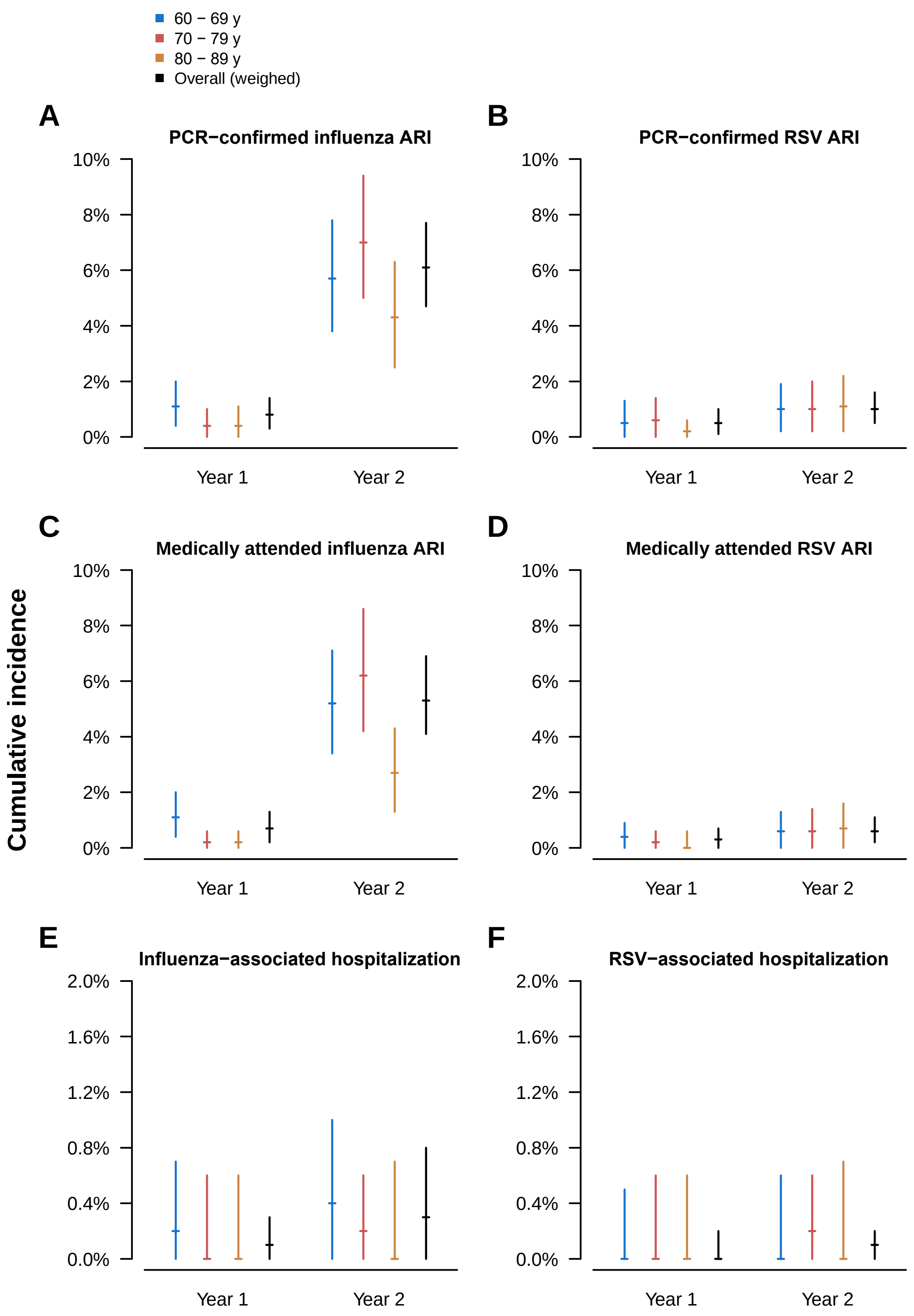

**Appendix Figure 3. Cumulative incidences of (A) PCR-confirmed influenza ARI, (B) PCR-confirmed RSV ARI, (C) medically attended influenza ARI, (D) medically attended RSV ARI, (E) influenza-associated hospitalization, and (F) RSV-associated hospitalization, by study year and age group.** Blue: 60–69 years old; Red: 70–79 years old; Orange: 80–89 years old; Black: All-ages and weighed with Jiangsu province population age and sex structure.

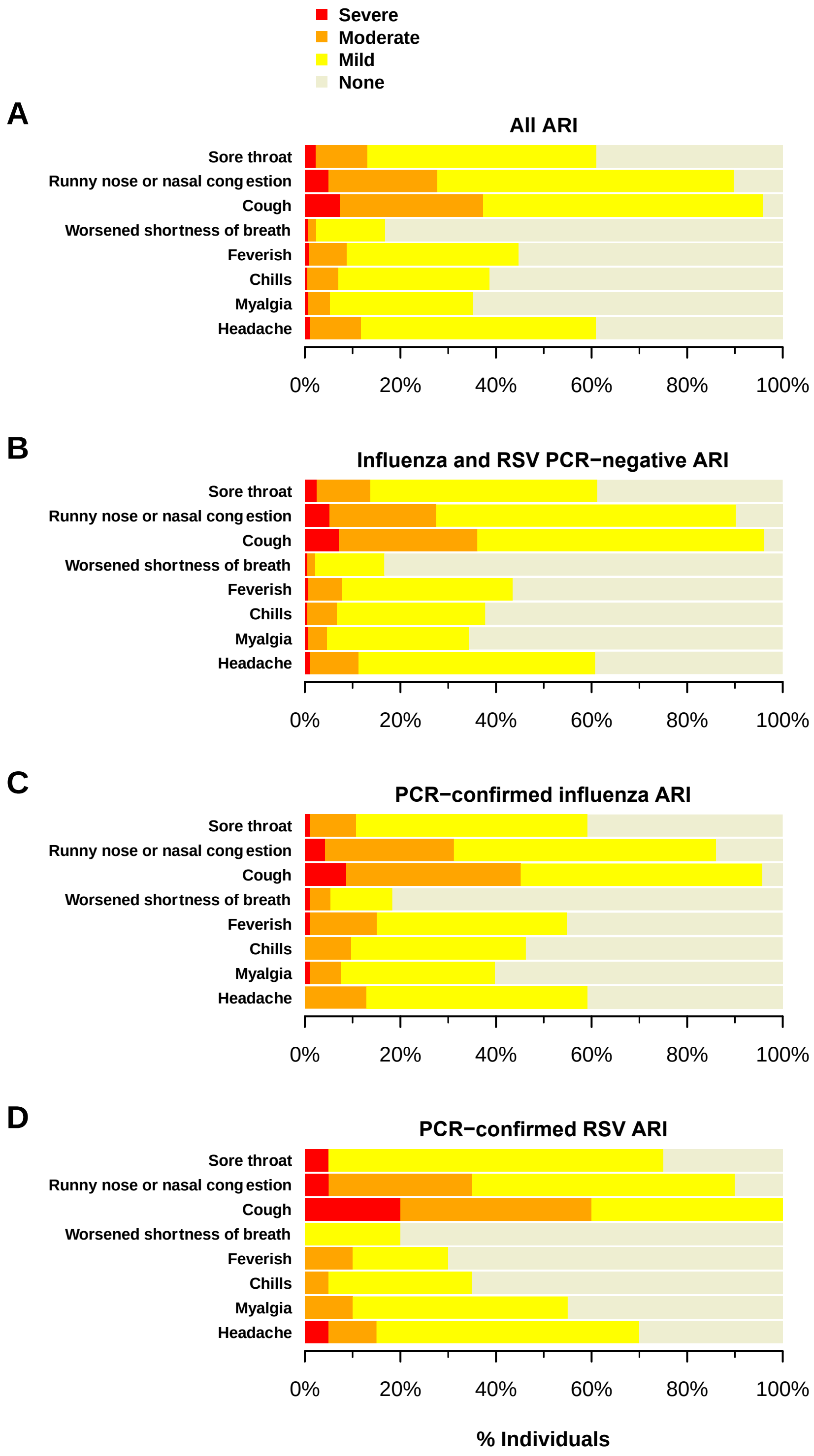

**Appendix Figure 4. Self-rated symptom severity for each symptom during ARI episodes among (A) all ARI, (B) influenza and RSV PCR-negative ARI, (C) PCR-confirmed influenza ARI and (D) PCR-confirmed RSV ARI.** Red: Severe; Orange: Moderate; Yellow: Mild; Grey: Not present.

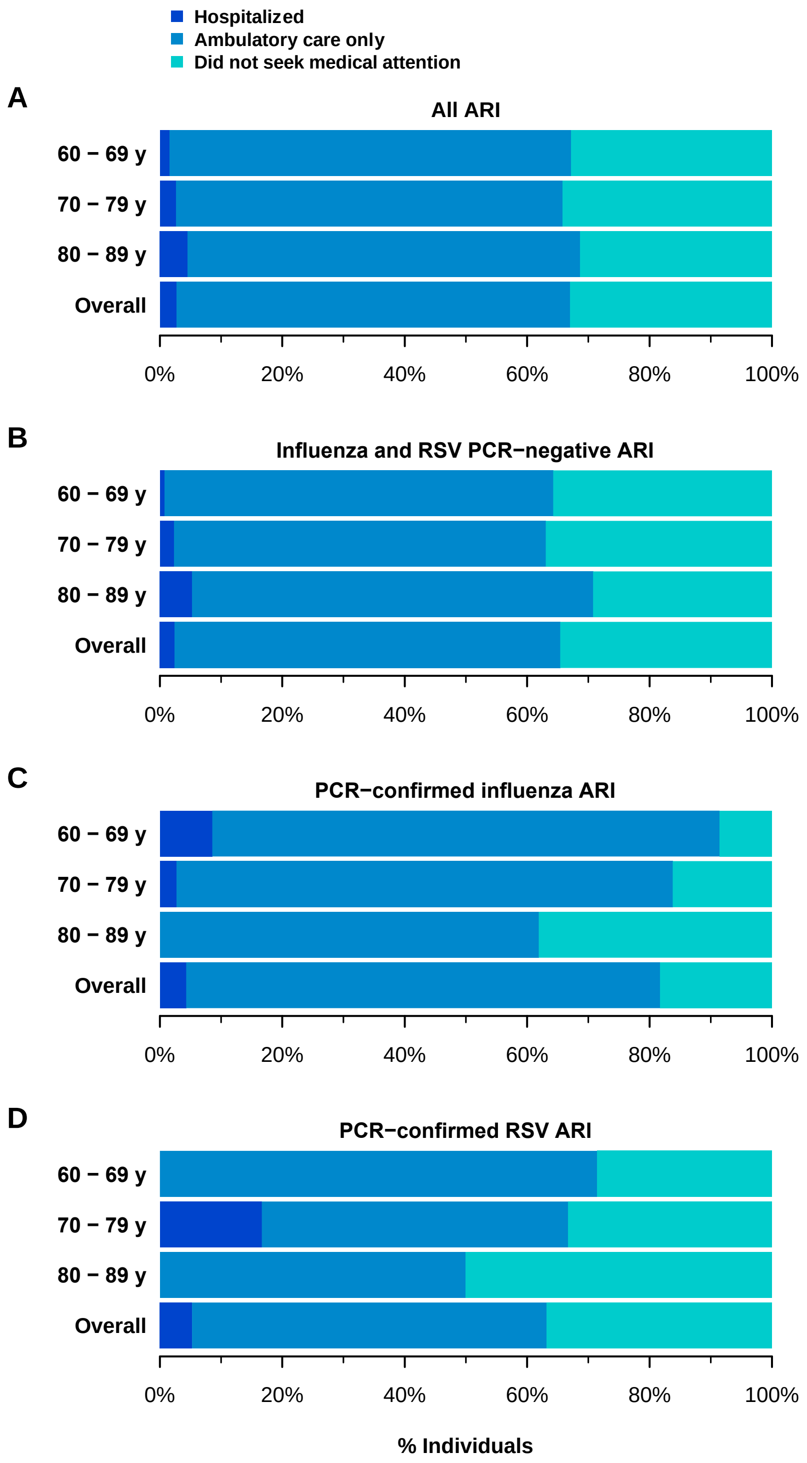

**Appendix Figure 5. Medical care utilization for ARI episodes by age group, among (A) all ARIs, (B) ARIs that were PCR-test negative for both influenza virus and RSV, (C) PCR-confirmed influenza ARIs and (D) PCR-confirmed RSV ARIs.** Dark blue: Participant was hospitalized; Light blue: Participant sought ambulatory care only (and was not hospitalized); Cyan: Participant did not seek medical attention.

### References

1. Yu H, Alonso WJ, Feng L, et al. Characterization of regional influenza seasonality patterns in China and implications for vaccination strategies: spatio-temporal modeling of surveillance data. *PLoS Med* 2013; **10**(11): e1001552.
2. Obando-Pacheco P, Justicia-Grande AJ, Rivero-Calle I, et al. Respiratory Syncytial Virus Seasonality: A Global Overview. *J Infect Dis* 2018; **217**(9): 1356-64.
3. Hayward AC, Fragaszy EB, Bermingham A, et al. Comparative community burden and severity of seasonal and pandemic influenza: results of the Flu Watch cohort study. *Lancet Respir Med* 2014; **2**(6): 445-54.
4. VanWormer JJ, Sundaram ME, Meece JK, Belongia EA. A cross-sectional analysis of symptom severity in adults with influenza and other acute respiratory illness in the outpatient setting. *BMC Infect Dis* 2014; **14**: 231.
5. Wu C, Gong Y, Wu J, et al. Chinese Version of the EQ-5D Preference Weights: Applicability in a Chinese General Population. *PLoS One* 2016; **11**(10): e0164334.
6. Borson S, Scanlan J, Brush M, Vitaliano P, Dokmak A. The mini-cog: a cognitive 'vital signs' measure for dementia screening in multi-lingual elderly. *Int J Geriatr Psychiatry* 2000; **15**(11): 1021-7.
7. Cowling BJ, Xu C, Tang F, et al. Cohort profile: the China Ageing REespiratory infections Study (CARES), a prospective cohort study in older adults in Eastern China. *BMJ Open* 2017; **7**(10): e017503.
8. Searle SD, Mitnitski A, Gahbauer EA, Gill TM, Rockwood K. A standard procedure for creating a frailty index. *BMC Geriatr* 2008; **8**: 24.
9. Harris PA, Taylor R, Thielke R, Payne J, Gonzalez N, Conde JG. Research electronic data capture (REDCap)--a metadata-driven methodology and workflow process for providing translational research informatics support. *J Biomed Inform* 2009; **42**(2): 377-81.
10. Fowler KB, Gupta V, Sullender W, et al. Incidence of symptomatic A(H1N1)pdm09 influenza during the pandemic and post-pandemic periods in a rural Indian community. *Int J Infect Dis* 2013; **17**(12): e1182-5.
11. Tinoco YO, Azziz-Baumgartner E, Uyeki TM, et al. Burden of Influenza in 4 Ecologically Distinct Regions of Peru: Household Active Surveillance of a Community Cohort, 2009-2015. *Clin Infect Dis* 2017; **65**(9): 1532-41.
12. National Notifiable Diseases Surveillance System (NNDSS). Downloads and Resources: MMWR Weeks Calendars. 2018. https://wwwn.cdc.gov/nndss/downloads.html (accessed October 18, 2016 2016).
13. Fowlkes A, Steffens A, Temte J, et al. Incidence of medically attended influenza during pandemic and post-pandemic seasons through the Influenza Incidence Surveillance Project, 2009-13. *Lancet Respir Med* 2015; **3**(9): 709-18.
14. Hanley JA, Lippman-Hand A. If nothing goes wrong, is everything all right? Interpreting zero numerators. *JAMA* 1983; **249**(13): 1743-5.
15. Cowling BJ, Chan KH, Fang VJ, et al. Comparative epidemiology of pandemic and seasonal influenza A in households. *N Engl J Med* 2010; **362**(23): 2175-84.
16. University of California. Global Administrative Areas. 2019. https://gadm.org/ (accessed October 21 2019).
